# Differentially Private Distributed Inference for Multicenter Clinical Studies

**DOI:** 10.1101/2025.03.10.25323686

**Authors:** Marios Papachristou, M. Amin Rahimian

## Abstract

Extracting reliable conclusions from data distributed across institutions is a core problem in healthcare, as pooling patient records across centers would improve inference; however, privacy regulations and the lack of a trusted central authority pose significant challenges that frequently delay or prevent multicenter studies. In this work, we develop a framework for differentially private distributed inference in which institutions repeatedly exchange log belief-ratio statistics with other participating institutions, subject to differential privacy (DP). We show that with arithmetic and geometric averaging of beliefs, we can control the false-negative and false-positive rates as functions of the privacy budget, communication rounds, and statistical separation between the hypotheses; exposing a three-way trade-off among accuracy, communication, and privacy. We derive finite-sample bounds on the Type I and Type II error probabilities that allow us to perform distributed hypothesis tests at a target significance level and show that the Laplace mechanism minimizes convergence time subject to differential privacy guarantees. For distributed online learning from data streams (e.g., in epidemiological surveillance or in studies with rolling recruitment), we show that privacy noise vanishes asymptotically and online learning admits similar finite-sample convergence guarantees. On simulated multicenter survival analyses on data from the AIDS Clinical Trials Group and an advanced-cancer cohort, and on a simulated genetic association study over New York City hospitals, our method approaches the non-private baseline at a small privacy budget between 1 and 10, and it runs 10× to 1000× faster than homomorphic-encryption methods, and incurs up to 100× lower error than first-order private convex optimization methods. Finally, the level of aggregation is a primary design choice: federating at the organizational rather than the hospital level simultaneously strengthens privacy, raises statistical power, and lowers communication and administrative burden, so the priority should be to pool data within organizations before setting up federated analytics frameworks.

## 1. Introduction

Clinical trials represent the gold standard for generating medical evidence and depend on recruiting a sufficient number of patients in order to perform valid inferences. However, patient recruitment, both in terms of the total number of patients as well as recruiting patients from underrepresented minorities, faces significant challenges. One of the most challenging obstacles that results in centers having lower sample sizes or samples that are not representative of the general population is patient privacy and regulatory compliance. The privacy risk is not hypothetical, even when only aggregate statistics are shared: from published allele frequencies alone, membership-inference attacks can determine whether a given individual contributed to a study (Homer et al. 2008); a finding that prompted the withdrawal of summary genomic data from open access and continues to impede collaboration (Zerhouni and Nabel 2008, Erlich and Narayanan 2014).

A potential solution to circumvent this problem is establishing multicenter studies where several participating institutions opt in to share their data, subject to data usage agreements (DUAs). Centralized trial networks such as the AIDS Clinical Trials Group or the UK Biobank have demonstrated the value of pooled multicenter recruitment (Hammer et al. 1996, Campbell et al. 2012, Sudlow et al. 2015), but they depend on coordination infrastructures that are beyond the operational capacity of many institutions, particularly smaller centers whose patient populations are often the most clinically relevant and demographically diverse.

Geographically separated institutions often hold patient data that, if jointly analyzed, would yield statistically reliable conclusions about treatment efficacy, prognostic factors, genetic associations, and disease mechanisms. Performing valid inferences collectively by pooling such data would therefore be of great value, were it not for the strict privacy regulations that constrain it. The downstream consequences of these regulations, namely HIPAA (1996) in the United States and GDPR (2018) in the European Union, are well-documented for multicenter collaborations (Ness 2007, Institute of Medicine 2009, Kao and Kang 2026, Clarke et al. 2019): such collaborations are routinely delayed or abandoned due to regulatory barriers, hindering discovery and innovation.

These barriers motivate the central question that this paper addresses:

*How can geographically distributed institutions jointly analyze sensitive patient data to support reliable statistical inference, without any institution relinquishing control of its own records or violating the confidentiality obligations it owes to its patients?*

We answer this question by developing a framework for differentially private distributed inference in which multiple centers exchange privacy-protected beliefs rather than patient records and perform social learning (Bullo et al. 2009, Jackson 2008, Golub and Sadler 2016, Rahimian and Jadbabaie 2016). Differential privacy (DP) provides the formal guarantee: a mechanism satisfying *ε*-DP ensures that the probability of any observable output changes by at most a factor of *e*^*ε*^ when any single patient’s data is added or removed, providing each patient with a quantifiable and formally verifiable form of plausible deniability (Dwork and Roth 2014). This guarantee is not merely contractual but mathematical, and it is auditable by regulators and patients alike (Dwork et al. 2019). By incorporating DP into iterative belief exchange protocols, our framework enables institutions to collectively reach statistically valid conclusions, such as performing hypothesis tests at a prescribed significance level, while mathematically preserving privacy and without the need for a centralized coordinating authority or specialized cryptographic infrastructure.

The practical relevance of this approach is underscored by recent policy developments. The U.S. Department of Health and Human Services’ Advancing Clinical Trial Readiness initiative, launched by ARPA-H (2024), seeks to establish a decentralized, on-demand clinical trial infrastructure accessible to 90% of eligible participants within thirty minutes of their home, automating data extraction and synchronization across electronic health records at distributed sites. Our framework addresses the core statistical challenge such an infrastructure must solve: how to extract reliable collective inference from distributed, privacy-constrained institutional data without centralized access to patient records.

### Main Contributions

We propose a differentially private distributed inference framework for information aggregation among a network of *n* institutions, each holding private patient data and engaging in iterative belief exchange to collectively identify the most likely alternatives (e.g., to choose from possible diagnoses or treatment options), test a hypothesis at a prescribed significance level (e.g., in genetic association and survival analyses), or learn the true state from a continuous stream of observations (e.g., for epidemiological surveillance). Privacy is enforced through calibrated Laplace noise injection into the belief statistics exchanged between neighboring institutions, ensuring that no institution’s communications reveal individual patient records.

We establish explicit finite-sample bounds on Type-I and Type-II error probabilities as functions of the privacy budget *ε*, the number of communication rounds *K*, the number of iterations per round *T*, and the statistical divergence between competing hypotheses, showing a three-way tradeoff among inference accuracy, communication complexity, and privacy level. To manage this tradeoff in practice, we develop two complementary aggregation strategies: arithmetic averaging, which controls Type-II error and is suited to screening contexts where missed detections carry the greater clinical cost, and geometric averaging, which controls Type-I error and is suited to regulatory contexts where false positives carry legal and clinical liability. A two-threshold algorithm provides simultaneous control over both error types at a modest increase in communication complexity. These guarantees extend naturally to formal hypothesis testing at a prescribed significance level *α*, and accommodate both simple and composite hypotheses including the generalized likelihood ratio tests used in survival analysis. For prospective multicenter trials and ongoing epidemiological surveillance, we further extend the framework to continuous observation streams, establishing that privacy noise vanishes asymptotically and providing finite-sample convergence guarantees characterizing the communication cost of privacy. In both the finite-sample and online settings, the Laplace mechanism is shown to be optimal for minimizing convergence time.

The most closely related prior work is Papachristou and Rahimian (2025), which incorporates differential privacy into the distributed learning framework of Rahimian and Jadbabaie (2016) for continuous parameter estimation, providing convergence guarantees in expectation on the *ℓ*_2_ norm of the estimation error. Our setting differs along three fundamental dimensions: inference in discrete parameter spaces introduces a failure probability bounded away from zero as iterations grow, requiring institutions to repeat deliberations across multiple rounds and aggregate results under an entirely new analytical framework; our finite-sample guarantees are expressed as explicit Type-I and Type-II error bounds rather than the expected *ℓ*_2_ error, the latter being insufficient to support hypothesis testing at a prescribed significance level; and the clinical applications we develop, multicenter survival analysis and differentially private genetic association analysis, are not addressed by Papachristou and Rahimian (2025) and require problem-specific sensitivity analyses and algorithmic adaptations. The optimality of the Laplace mechanism for minimizing convergence time, which also arises in the continuous-estimation setting of Papachristou and Rahimian (2025), thus persists across two structurally distinct inference settings, providing independent validation of this design choice.

The complementary works of Rizk et al. (2023) and Cyffers et al. (2024) provide first-order methods for distributed optimization and learning subject to DP. Cyffers et al. (2024) analyzes the random-walk first-order stochastic gradient descent method under pairwise network DP. Additionally, Rizk et al. (2023) provides consensus algorithms and introduces the graph-homomorphic noise model to achieve DP. We note that both Cyffers et al. (2024) and Rizk et al. (2023), provide convergence guarantees to the optimum compared to our method, which gives Type-I and Type-II error guarantees. Later in the paper, we show that our method achieves significantly lower error than first-order methods, as our methods rely on adding noise only once to the statistics before propagation, where, in contrast, first-order methods need to repeatedly add noise to the gradient.

We validate the framework on three healthcare applications of increasing scale and complexity: multicenter survival analysis on data from ACTG Study 175 under the Cox proportional hazards model (Cox 1972); survival analysis of advanced cancer patients from Samstein et al. (2019) assessing the prognostic value of tumor mutational burden; and a large-scale simulation of differentially private genetic region study on a federated network derived from the New York City hospital system, comprising 84 hospitals across 16 parent organizations with *N* = 25,000 simulated individuals. Our framework is able to achieve statistical performance approaching the centralized baseline under practically deployable privacy budgets, at runtimes between 10× and 1,000× faster than encryption-based baselines (Froelicher et al. 2021, Geva et al. 2023), and with substantially lower error than existing first-order differentially private optimization methods (Rizk et al. 2023). Analysis of the New York City hospital network further reveals that organizational-level federation simultaneously dominates hospital-level federation in privacy strength, statistical power, and communication complexity providing a principled quantitative basis for governance decisions which, historically, are made without formal analytical grounding, with concrete managerial implications.

### 1.1 Motivating Example: Federated Survival Analysis for Multicenter Clinical Trials

Survival analysis is widely used to examine how patients’ survival outcomes (i.e., timing of events such as death, relapse, remission, recovery, etc.) vary in response to specific treatments or other health factors. Importantly, survival analysis accounts for censoring, which refers to the existence of data points where events of interest, such as death, hospitalization, or remission, have not occurred by the end of the study period, yet these data points are informative about the effect of treatments. Survival models account for censoring to give more accurate and unbiased estimates. This approach typically involves key metrics such as survival functions, hazard rates, and median survival times, allowing researchers to compare the effectiveness of different treatments or identify prognostic factors. Standard nonparametric and semiparametric models, such as the Kaplan-Meier curve (Kaplan and Meier 1958) and the Cox proportional hazards model (Cox 1972), allow us to analyze patient survival data with great flexibility in various clinical and experimental settings. By modeling time-to-event data, survival analysis plays a crucial role in clinical trials, epidemiological studies, and personalized medicine, helping clinicians and researchers make informed decisions about treatment strategies and patient care. Our simulation study in Section 4 consists of survival analysis on a data set from the AIDS Clinical Trials Group (ACTG) network (National Institute of Allergy and Infectious Diseases 1995), which has conducted several studies of HIV-infected and AIDS patients since the late 1980s (Hammer et al. 1996, Campbell et al. 2012).

Healthcare centers vary in the demographics of the patients that they serve, and they may even have different target groups (e.g., children’s or women’s hospitals). Although different centers can conduct clinical trials and hypothesis tests on their own, potential data limitations can yield false conclusions, for example, due to small sample sizes or higher prevalence of specific health conditions among some demographics that are under- or overrepresented in their patient population. For this reason, multiple healthcare centers can join multicenter trials to perform hypothesis tests with more accuracy, using larger and more diverse patient samples. To model multicenter trials, we divide patient data from the ACTG study 175 (National Institute of Allergy and Infectious Diseases 1995) equally at random between five centers and use the proportional hazards model (Cox 1972) for survival analysis. For ease of exposition, we consider a simple hypothesis testing scenario to determine the efficacy and safety of a new treatment (ddI) against standard care (ZDV), which are antiretroviral HIV / AIDS medications in the ACTG study 175. For each center *i* ∈ [*n*], we denote their data set of *n*_*i*_ patients by ***s***_*i*_. The patient *j* in the center *i* has the treatment variable ***x***_*i j*_ ∈ {0, 1} which indicates if they have received the new treatment (ddI), ***x***_*i j*_ = 1, or standard care (ZDV), ***x***_*i j*_ = 0, and control variables ***z***_*i j*_ . The effect of the treatment is parametrized by *θ*_trt_. The survival event for each patient is denoted by ***δ***_*i j*_ ∈ {0, 1} with ***δ***_*i j*_ = 1 corresponding to death (survival time is observed) and ***δ***_*i j*_ = 0 corresponding to survival (survival time is censored). The survival or censoring time is measured in days and is indicated by ***t***_*i j*_ ∈ ℕ. Following the Cox proportional hazards model, the partial log-likelihood of the patient survival data for center *i* is given by:

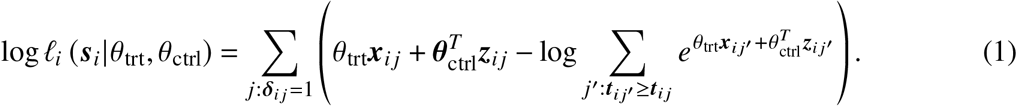

Note that using *ℓ*_*i*_ in (1) is with some abuse of notation because it does not represent the full likelihood of the observed data; however, under Cox’s proportional hazards model we can reliably infer the effect of the new treatment and other risk factors that can be included as covariates along with the treatment variable in Equation (1), from the partial likelihoods without specifying the full likelihoods or the functional form of the baseline hazard rate corresponding to ***x***_*i j*_ = 0. We are interested in rejecting the simple null hypothesis that *θ*_trt_ = 0, i.e., that the two survival curves are the same, which, in the absence of other covariates, is equivalent to a nonparametric log-rank test of survival times between the treatment and control groups. For concreteness, we use *θ*_trt_ = *θ*_1_ = − log(2) as the simple alternative, which corresponds to patients being twice as likely to survive under the new treatment than in standard care (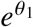is the relative risk for the new treatment under the proportional hazards model).

Figure 1(a) shows that the sample size of Kaplan-Meier survival curves calculated by one of the hospitals is not large enough to detect a difference between the two treatments, and, in fact, fitting the proportional hazards model using data from one hospital yields a *p*-value of 0.11 in Figure 1(c). However, if centers were to pool their data (ignoring privacy concerns) through a centralized authority, then the centralized authority could perform the test at a larger sample size. Figure 1(b) shows the Kaplan-Meier survival curves are easily distinguishable in the centralized case. In Figure 1(c), when all data are pooled together, the difference between log-hazard ratios (log HR) from fitting proportional hazards models for the two treatments is statistically significant at *p* < 0.001.

**Figure 1.**
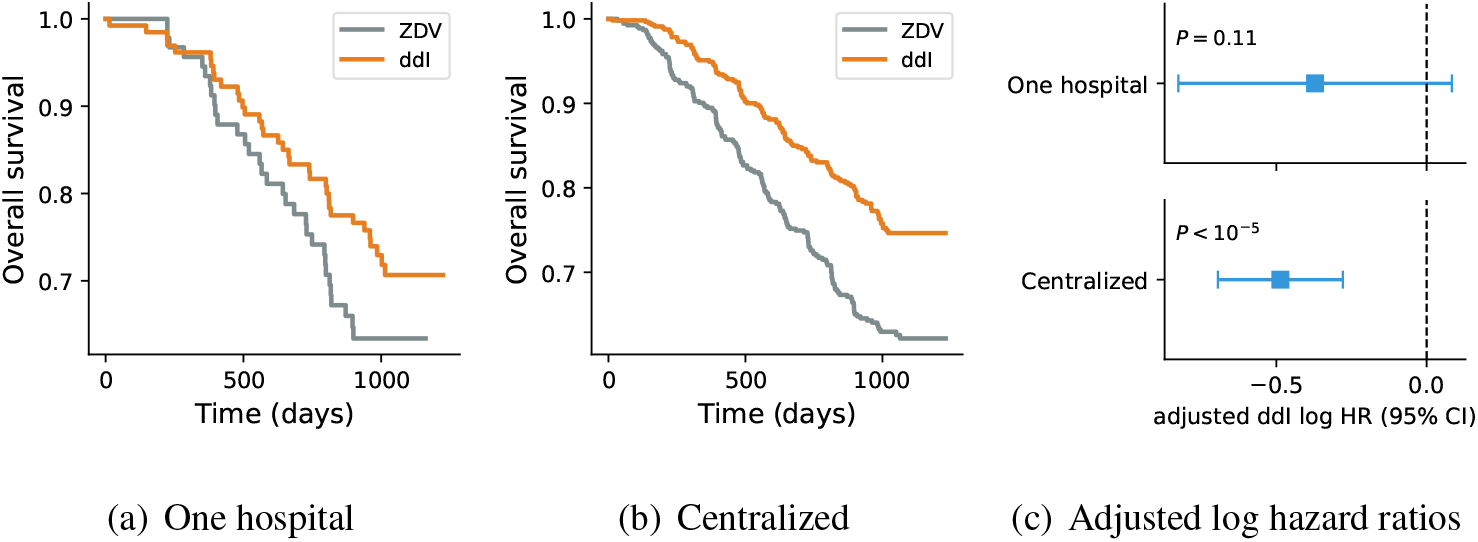
Survival analysis for ACTG study 175 (National Institute of Allergy and Infectious Diseases 1995) comparing the ZDV and ddI treatments. ZDV stands for zidovudine, and ddI stands for didanosine, and the data is split equally among five hospitals. **Left and middle:** Kaplan-Meier survival curves for one hospital and for all the data pooled (centralized). **Right:** ddI log hazard ratios with 95% confidence intervals from a proportional hazards model adjusted for baseline covariates (age, CD4 count, Karnofsky score, weight, prior antiretroviral exposure, race, and sex), fitted on all the data (centralized) and on one hospital. A single hospital cannot resolve the effect whereas the pooled analysis is highly significant.

In a distributed setting where privacy concerns limit data sharing and preclude data pooling for centralized access, centers can exchange noisy information locally to achieve distributed hypothesis testing with privacy guarantees. In the private regime, inspired by the likelihood ratio test, we propose that each center calculate its local (partial) log-likelihood ratio statistic log 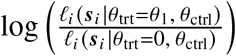, add appropriately chosen Laplace noise ***d***_*i*_, and then exchange the noisy statistic with its neighbors; see the simulation study in Section 4. The methods we devise in Sections 2 and 3 allow centers to form belief statistics locally, in a privacy-preserving manner, and use them for collective hypothesis testing with guarantees of false positive and false negative rates.

## 2. Belief Propagation for Differentially Private Distributed Inference

More generally, each center can have access to private observations with different likelihoods (for example, representing different populations of patients in different centers), which are parametrized by a common, finite set of alternatives. The goal of the centers is to exchange information to calculate the likelihoods of their collective observations and to choose a common set of maximum likelihood estimates (MLEs). To this end, centers exchange (non-Bayesian) belief statistics and decide on a common set of best alternatives (collective likelihood maximizers) based on their convergent beliefs. This MLE setup has natural extensions to hypothesis testing and online learning that we explore in Section 3.2 and Appendix F, respectively. Before focusing on the theoretical performance and privacy guarantees of our distributed belief exchange algorithms in Section 3, we introduce our information environment and the setup of distributed inference in Section 2.1. Our belief exchange rules have a log-linear format, which has normative foundations in decision analysis (Abbas 2009), and can also be justified on the basis of their convergence properties (Olshevsky 2017). Due to length constraints, we elaborate on these connections in Appendix C.

### 2.1 Problem Formulation: Distributed Inference & Learning in Discrete Spaces

A collection of *n* centers, denoted by the set [*n*] = {1, …, *n*}, wants to select an alternative from a finite set Θ. Each center *i* has its own data, which are fixed at the beginning or arrive in an online stream over time. The data follow a parametric model given by its likelihood function *ℓ*_*i*_ (·|*θ*), *θ* ∈ Θ. The goal of the centers is to exchange information to select a set of alternatives that best describes their data collectively, in a maximum likelihood sense. Information exchange between centers (e.g., collaborating hospitals) is constrained by organizational and legal barriers, as well as *privacy regulations* (e.g., HIPAA or GDPR) that limit who can exchange what information with whom. In our framework, organizational and legal barriers are captured by the structure of the communication network that limits information exchange to permissible local neighborhoods, and protection of private data (e.g., protected health information at each hospital) is achieved through differential privacy.

Centers are connected according to an undirected graph *G*([*n*], ℰ) on *n* nodes indexed by [*n*] with an edge set ℰ that contains no self-loops. The neighborhood of center *i* in *G* is denoted by *N*_*i*_ and corresponds to all centers with whom center *i* can exchange information locally. The graph is associated with an irreducible and aperiodic (primitive), doubly stochastic adjacency matrix *A* = [*a*_*ij*_]_*i, j* ∈[*n*]_ with weights *a*_*i j*_ = 0 whenever (*i, j*) ∉ ℰ and weight *a*_*i j*_ > 0 for all (*i, j*) ∈ ℰ, and moreover, ∑ _*i*∈[*n*]_ *a*_*i j*_ = _*j*∈[*n*]_ *a*_*i j*_ = 1. The *n* eigenvalues of the adjacency matrix *A* are ordered by their modulus and denoted by 0 < |*λ*_*n*_ (*A*) | ≤ · · · ≤ |*λ*_2_(*A*) | < *λ*_1_(*A*) = 1, with their associated set of bi-orthonormal eigenvectors, {***l***_*i*_, ***r***_*i*_ }_*i*∈[*n*]_, satisfying 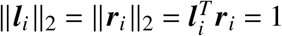for all *i* ∈ [*n*] and 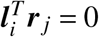for all *i* ≠ *j* . We use E [·] and V [·] to denote expectation and variance operators.

#### The Privacy Notion

Differential privacy restrictions dictate the protection of individual data, that is, ensuring that information exchange mechanisms between centers satisfy the following criteria for *ε*-DP.

##### Definition 1.

A mechanism ℳ_*i*_ : *S* → ℛ, acting on private patient datasets (or signals), is said to be *ε*-DP with respect to the patient dataset ***s*** ∈ *S* ⊂ ℝ^*d*^ if and only if for all *X* ⊆ ℛ we have that ℙ[ℳ_*i*_ (***s***) ∈ *X*] ≤ *e*^*ε*^ℙ[ℳ_*i*_ (***s***^′^) ∈ *X*] for any pair of adjacent patient datasets ***s*** ∼ ***s***^′^ that differ in at most one patient’s data.

The *ε*-DP constraint on ℳ_*i*_ ensures that as *ε* → 0, no information can be inferred about whether ***s*** or any of its adjacent points (i.e., ***s***^′^ ∈ *S* : ***s***^′^ ∼ ***s***) are the input that generates the observed output ℳ_*i*_ (***s***). Our primary interest in this work is in belief exchange mechanisms for which the range space ℛ is the probability simplex of all distributions over the state space Θ, denoted by Simplex(Θ). Defining the appropriate notion of adjacency allows us to control sensitive information leaks.

#### The Distributed Inference Problem

We consider a distributed information environment, where the goal is to estimate the best alternative or perform a hypothesis test in a distributed manner based on a set of initial observations. In Appendix F, we consider an online learning extension where the goal is to learn the true alternative online by repeatedly observing data streams over time. In multicenter studies, the online learning setup corresponds to centers recruiting patients over time. The centers can rely on each other’s observational capabilities to recruit a diverse patient demographic, which they may otherwise lack access to, compromising their ability to detect effects in certain populations.

In the distributed maximum likelihood estimation task (MLE), there is a set of likelihood maximizers Θ^★^ ⊂ Θ, and centers aim to determine Θ^★^ collectively by combining their private patient datasets while communicating within their local neighborhoods. Specifically, each center has access to a dataset ***s***_*i*_ ∈ *S* of *n*_*i*_ patients that obeys a likelihood function 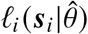for all 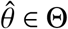. The centers’ task is, given patient datasets ***s***_*i*_ for each center *i* ∈ [*n*], to find the set of likelihood maximizers, i.e.,

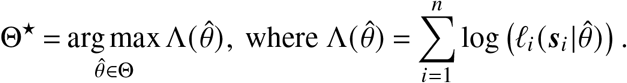

We define 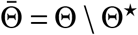as the set of non-optimal states. We let *f* ^★^ represent the proportion of MLE states, i.e., *f* ^★^ = |Θ^★^|/|Θ|. Rahimian and Jadbabaie (2016) give a non-private algorithm for belief exchange, described in Appendix D, that is able to recover Θ^★^ asymptotically.

In the multicenter clinical trial example of Section 1.1, the state space is Θ = {0, *θ*_1_}, where *θ* = 0 corresponds to the null hypothesis that the two treatments are statistically indistinguishable, and *θ* = *θ*_1_ < 0 corresponds to the hypothesis that ddI improves patient survival compared to ZDV, as is the case in Figure 1; hence, Θ^★^ = {*θ*_1_}. The goal of the centers is to collectively infer that *θ* = *θ*_1_ is the best state (MLE) given the data of the patients because Λ(*θ*_1_) > Λ(0). In Section 3.2, we show how distributed MLE algorithms can be modified for distributed hypothesis testing with statistical significance guarantees.

### 2.2 Performance Analysis Framework

Our proposed log-linear update rules rely on adding zero-mean Laplace noise to the log-likelihood statistics of the private data. Ideally, by adding zero-mean noise in the MLE problem, we would anticipate that log-linear updates should recover MLE states Θ^★^ in expectation. However, the added noise makes our algorithm output random, so we need a systematic way to control the error of the output and achieve a desired accuracy level by repeating the algorithm. Thus, providing high-probability guarantees for our algorithms comes at a cost: The algorithms need to be repeated in sufficiently many rounds, and their outputs combined across these rounds.

#### Type I and Type II error rates

To control the output uncertainty resulting from privacy-preserving randomization noise, our distributed MLE algorithms, in addition to respecting a set DP budget *ε* > 0, also admit two types of error guarantees, which we refer to as Type I and Type II error probabilities and denote by *α* and 1 − *β*, following the hypothesis testing nomenclature, where *β* denotes the power of the test. In fact, in Section 3.2 (with proofs and convergence analysis in Appendix H), we show that our distributed MLE algorithms and guarantees can be adapted to perform a distributed hypothesis test at the significance level *α*. For a distributed MLE algorithm *A* that returns a set of maximum likelihood estimators 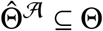, we define the *Type I* error rate *α* as the probability of the event that 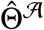includes non-MLE states, i.e., 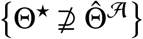, which can be controlled by bounding 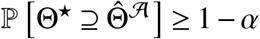. Similarly, we define the *Type II* errorrate 1 − *β* as the probability of the event that 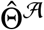fails to include some MLE states, i.e., 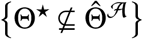, which can be controlled by bounding 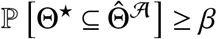. A Type I error occurs whenever 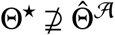, for example, when the algorithm outputs too many states, including some that do not belong to Θ^★^. A Type II error occurs whenever 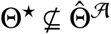, for example, when the algorithm filters too many states, missing some MLE states in its output.

Returning to our examples from Section 1, when selecting among treatments with severe side effects, we want to limit the possibility of selecting ineffective treatments by ensuring a small Type I error rate *α* so that 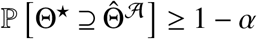. Similarly, when conducting a clinical trial or a genome-wide association study, we need to ensure the efficacy of treatments in clinical trials or the discovery of significant single nucleotide polymorphisms (SNPs) associations at a desired level of statistical significance (e.g., *α* = 0.05). On the other hand, when developing a new cancer screening tool, we want MLE states to be included in the algorithm output 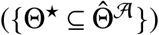with high probability, that is, for *β* to be high (e.g., *β* = 0.8). This will ensure that we can detect all patients who are at high risk of cancer (Θ^★^). In this case, it is important to control the Type II error rate so that 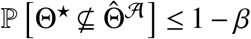for a large enough *β*, but it may be acceptable to misidentify some patients as high risk (i.e., for Θ^*A*^ to include some non-MLE states so that 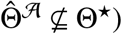. In Section 3.1, we show that arithmetic or geometric averaging of beliefs across the rounds may each be suitable, depending on the type of error that one needs to preclude or control.

#### Communication complexity

To achieve guarantees on the t wo error probabilities, *α* and 1 − *β*, iterations in each round, where *η* corresponds to either *α* or 1 − *β*, subject to a DP budget *ε*, we must run our algorithms in *K* (*ε, η*, Θ, *G*, {*ℓ*_*i*_ (·|·)}_*i*∈[*n*]_) rounds and with *T* (*ε, η*, Θ, *G*, {*ℓ*_*i*_ (·|·)}_*i*∈[*n*]_) or the maximum of the two (depending on the algorithm and the error controls that it provides). We define

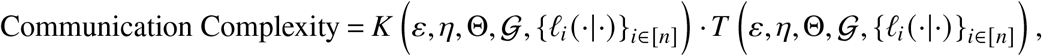

as the total number of belief updates. Beyond the privacy budget *ε*, the error probability *η*, the number of centers *n*, and the algorithm design parameters, the communication complexity bounds (see also Appendix B for a summary of the results) depend on the following statistical properties of the information environment:

- ***Private signal structures:*** In the MLE case, our bounds depend on the largest log-likelihood magnitude:

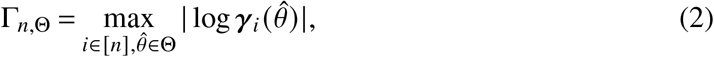

the minimum divergence between any non-MLE state and an MLE state:

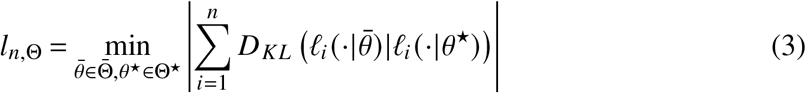

##### Algorithm 1

Private Distributed MLE (AM/GM)

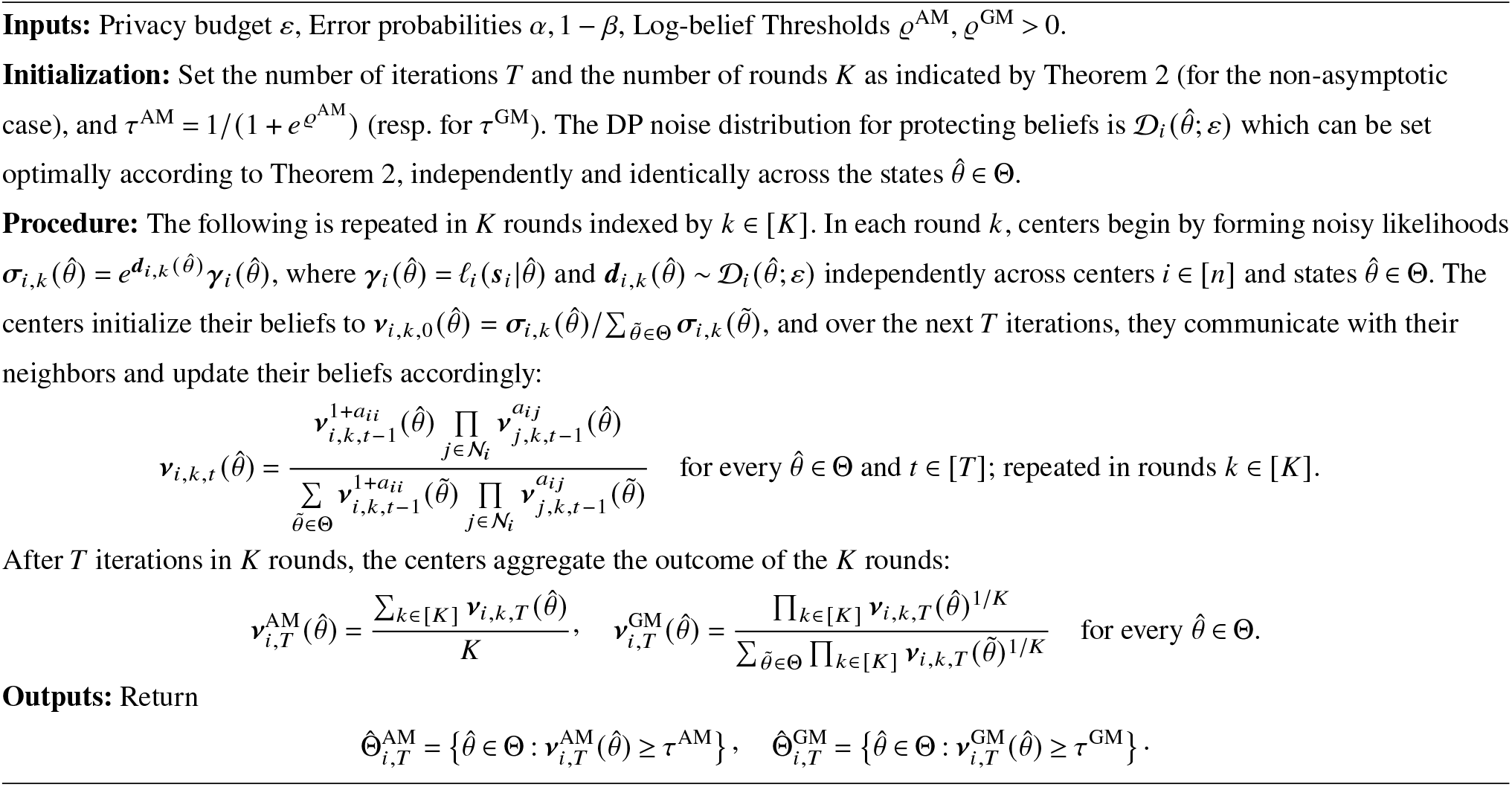

and the maximum standard deviation of the log-likelihood ratio statistics:

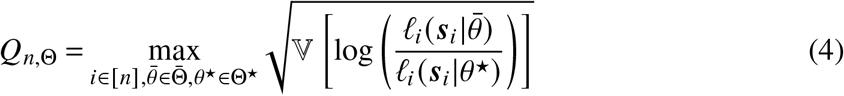

- ***Sensitivity:*** To satisfy the *ε*-DP requirement in Definition 1, the noise ***d***_*i*_ can be set according to the Laplace mechanism (Dwork and Roth 2014, Section 3.3), which adds Laplace distribution noise in the log space: 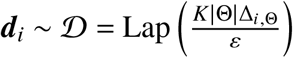, where *K* |Θ|Δ_*i*,Θ_/*ε* is the scale parameter of the Laplace distribution and the global *ℓ*_1_-sensitivity, Δ_*i*,Θ_, is defined as:

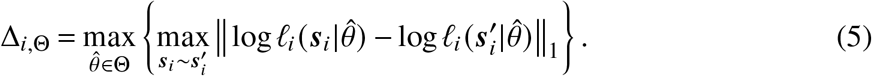

Intuitively, when log-likelihoods are highly sensitive to signal values more noise is required to mask the input signals at the *ε*-DP level. Here we have used *ε*/(*K* |Θ|) privacy budget per state per round to ensure that the initial beliefs are *ε*-DP overall by the composition property of DP (Dwork and Roth 2014, Theorem 3.14), after computing its value at all states 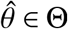. We state the non asymptotic bounds in terms of the maximum of the sensitivities 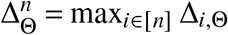.

## 3. Performance and Privacy Guarantees for Distributed Inference

### 3.1. Differentially Private Distributed MLE

The first idea behind the MLE algorithm is to introduce multiplicative noise subject to DP to the likelihood functions 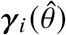. Multiplicative noise, which corresponds to additive noise in the log domain, ensures privacy. The centers then communicate their beliefs about the states with their local neighbors and form estimates of the true MLE over time. However, introducing noise makes the hypothesis selection randomized; therefore, the centers may misidentify the MLE states with a non-zero probability. To overcome this problem and guarantee Type I (resp. Type II) error at most *α* (resp. 1 − *β*), we run the algorithm for *K* independent rounds and combine these *K* estimates to produce the final beliefs. In Theorem 1, we show how an appropriate choice of *K* allows us to control the algorithm’s errors.

To produce the final estimates, we rely on two ways of aggregating the *K* rounds of the algorithm: The first approach is to take the arithmetic mean of the *K* rounds to produce the final belief, which we call the AM estimator. The benefit of the AM estimator is that it can accurately, with high probability, identify all the MLE states; i.e., we can control its Type II error probability: 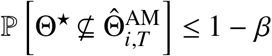for all centers *i* ∈ [*n*] and *K* and *T* large enough. This estimator has high recall but low precision because it can recover a superset of Θ^★^. The second way to combine the *K* rounds is to take the geometric mean, which we call the GM estimator. For the GM estimator, we can control the Type I error probability: 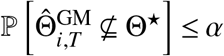, by choosing *K* and *T* large enough to ensure that it recovers a subset of Θ^★^ with high probability. We first start with a result on the asymptotic behavior of Algorithm 1. Specifically, we show that we can recover the MLE with high probability by repeating the algorithm *K* times for appropriately chosen *K* (proved in Appendix G.2):

#### Theorem 1.

*Consider Algorithm 1 run with state-independent noise distributions* 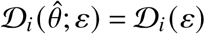*that satisfy ε-DP for a fixed ε* > 0. *Then, for every center i* ∈ [*n*], *the following ho ld:*

- ***Type II error (AM estimator):*** *if* 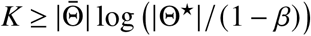, *then* 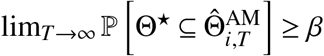*moreover*, 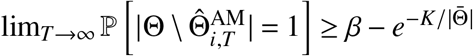.
- ***Type I error (GM estimator):*** *if* 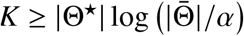, *then* 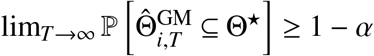*moreover*,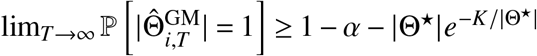.
- *The beliefs exchanged and the resulting estimates are ε-DP with respect to private signals*.

Repeating Algorithm 1 for *K* ≥ |Θ| log(|Θ|/min{*α*, 1 − *β*}) iterations, one can assert 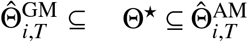as *T* → ∞ with probability *β* − *α* without knowing *f* ^★^. Also, it is interesting to point out that the above theorem holds regardless of the noise distribution, as long as the noise distribution does not depend on the state for a given center and it satisfies *ε*-DP.

In practice, the hyperparameters *K* and *T* can be set optimally using Theorem 2, which provides explicit, closed-form expressions for both. Moreover, for the common case of hypothesis testing with two states, the GM estimators recover the correct hypothesis exactly, as confirmed by our theoretical and empirical results in Sections 3.2 and 4, respectively. For richer state spaces, our two-threshold algorithm (deferred to Appendix E) enables simultaneous control of both Type-I and Type-II errors without this tension. Nevertheless, it is instructive to understand the role of *K* in the AM/GM setting: the error probability decreases exponentially in *K*, while the noise standard deviation scales linearly, creating a bias–variance-like trade-off with increasing *K*. This is reflected in Theorem 1: as *K, T* → ∞, the GM estimator concentrates on a singleton (high precision, potentially lower recall), while the AM estimator expands toward the full set (high recall, potentially lower precision). Choosing *K* as prescribed by Theorem 2 optimally balances these effects and yields the target error rates *α* and 1 − *β*.

The distributed MLE algorithm is repeated *K* times for each of the |Θ| states, and an attacker would have more observations of the outputs from the same signals; therefore, we need to rescale the privacy budget by *K* |Θ| to account for the number of runs and states. Thus, choosing 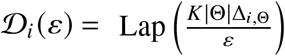satisfies *ε*-DP and the assumptions of Theorem 1. Our nonasymptotic analysis (presented next) shows that 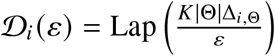is indeed optimal to minimize the required number of iterations per round.

The previous analysis focused on the regime where *T* → ∞. The next question that arises is to study the behavior of Algorithm 1 for finite *T*. As expected, the speed of convergence depends on the sum of standard deviations of the noise, that is, 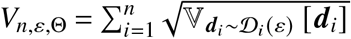. Our main result for private distributed MLE using the AM/GM method follows (proved in Appendix G.3):

#### Theorem 2.

*Let* 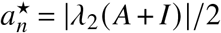*be the second-largest eigenvalue modulus (SLEM) of* (*A*+ *I*)/2. *For Algorithm 1 with thresholds ϱ*^AM^, *ϱ*^GM^ > 0 *and privacy budget ε* > 0, *the following hold for every center i* ∈ [*n*]:

- *For* 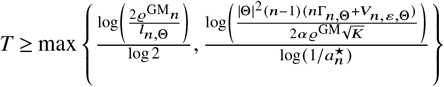*and* 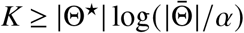, *we have* 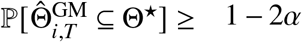. *Moreover*, 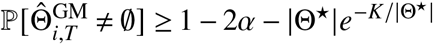.
- *For* 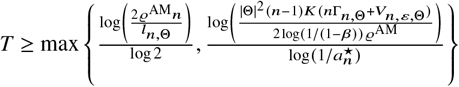*and* 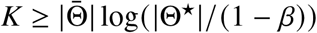, *we have* 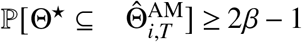. *Moreover*, 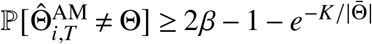.
- *The noise distributions that minimize the convergence time T are the Laplace distributions* 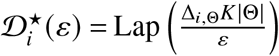.
- *The beliefs exchanged and the resulting estimates are ε-DP with respect to private signals*.

Recall that Γ_*n*,Θ_ and *l*_*n*,Θ_ that appear in the communication complexity bounds in Theorem 2 are properties of the signal structure defined in Equations (2) and (3). To minimize communication complexity, it suffices to pick the optimal threshold that makes the terms inside the maximum equal.

#### A Two-threshold Algorithm for Distributed MLE

Beyond the AM/GM algorithm, we develop a two-threshold algorithm that enables centers to jointly control both Type I and Type II errors in distributed maximum-likelihood estimation while preserving privacy. The method tracks how often each hypothesis’ belief exceeds two confidence thresholds across multiple communication rounds, effectively distinguishing MLE from non-MLE states. We show that, with appropriately chosen thresholds and a sufficient number of communication rounds, the algorithm identifies all MLE states while excluding non-MLE ones with high probability, achieving asymptotic recovery of the correct hypothesis set. Compared to single-threshold or averaging methods, the two-threshold approach offers finer control over false positive and false negative rates at a modest increase in communication complexity, as formalized in our asymptotic and non-asymptotic analyses. Due to length constraints, we defer the discussion of the two-threshold algorithm to Appendix E. Appendix B presents a summary of the main results obtained from the two-threshold algorithm.

### 3.2 Differentially Private Distributed Hypothesis Testing

The results we devised above for the MLE have a natural application in the case of hypothesis testing with simple or composite hypotheses. Specifically, we can specialize our designs to decentralized hypothesis testing at a significance level *α* as follows: We run the GM algorithm for *T* iterations and *K* rounds and set a threshold *ϱ*_*d*_ to reject the null hypothesis if the log-belief ratio exceeds it: 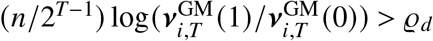. For the case of a simple null and a simple alternative, the threshold *ϱ*_*d*_ can be derived from the threshold *ϱ*_*c*_ of the corresponding uniformly most powerful (UMP) centralized log-likelihood ratio test (LRT) at level *α*/2. For sufficiently large *T* and *K*, we can show that

#### Proposition 1 (Simple Hypothesis Testing)

*Let α* ∈ (0, 1) *be a significance level. Let ϱ*_*c*_ *be the threshold of the UMP centralized log-likelihood ratio test at level α*/2, *such that* ℙ[2(Λ(1) −Λ(0)) ≥ *ϱ*_*c*_ |*θ* = 0] = *α*/2. *Run the GM algorithm (Algorithm 1) with a Type I error guarantee α*/2, *with T and K as in Theorem 2 and the rejection threshold ϱ*_*d*_ = *ϱ*_*c*_ − 1. *Then the decentralized test controls the Type I error at level α, that is*,

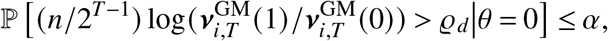

*and the resulting test is ε-DP*.

The idea behind the result is that, in practice, for sufficiently large *T* and *K*, 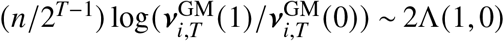with high probability (with probability at least 1 − *α*/2 due to Chebyshev’s inequality). The formal proof of Proposition 1 is in Appendix H.1.

However, in general, when the distribution of the centralized statistic under the null is not available, we cannot choose *ϱ*_*c*_ based on the UMP centralized LRT. In that case, the centers can perform a generalized likelihood ratio test locally and propagate the statistic. This sacrifices the optimality of the UMP test; however, it is more practical in real-world scenarios where the distribution of the sum of the statistics may be unknown under the null hypothesis.

Specifically, the generalized likelihood ratio test can be used to test a composite null hypothesis 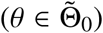against a composite alternative 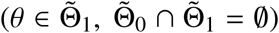, and requires a log-likelihood function that is twice differentiable and defined in a continuous parameter space 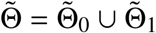.Specifically, in that case, each center initializes their belief using 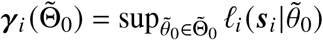and 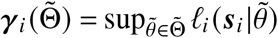, and runs the GM algorithm with the binary (meta) state space 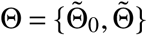. By Wilks’ theorem (Casella and Berger 2002, Theorem 10.3.1), for a large number of patients *n*_*i*_, each local log-likelihood ratio statistic is asymptotically distributed according to the 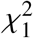distribution, and thus the sum of independent local statistics of the *n* centers follows a 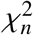distribution. We can use the asymptotic distribution of the sum of the local log-likelihood ratio statistics to set the rejection threshold for a distributed level *α* test to be 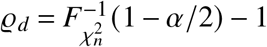. In contrast, the centralized test has a threshold of 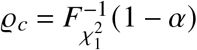. We provide additional details for the extension to composite hypothesis tests in Appendix H.2.

#### Extensions to Online Learning from Intermittent Streams

For prospective multicenter trials and epidemiological surveillance where centers recruit patients continuously over time, we extend the framework to online learning from intermittent data streams. Unlike the MLE setting, the online setting does not require repeating the algorithm across multiple rounds because the effect of DP noise vanishes asymptotically as a consequence of the Cesáro mean and the weak law of large numbers. We provide finite-sample convergence guarantees characterizing the communication cost of privacy and show that the Laplace mechanism remains optimal for minimizing convergence time. Due to space considerations, the algorithm and its analysis are presented in Appendix F.

## 4. Real-world Healthcare Data Sharing Applications

We validate the proposed framework on three healthcare applications of increasing scale and institutional complexity. The first two applications involve multicenter survival analysis on realworld clinical trial data: ACTG Study 175, evaluating the comparative efficacy of antiretroviral treatments on patient survival under the Cox proportional hazards model (Cox 1972), and a study of advanced cancer patients from Samstein et al. (2019), assessing tumor mutational burden as a prognostic factor under the immune checkpoint inhibitor therapy. The third application is a large-scale differentially private genetic region study instantiated on a federated network derived from the New York City hospital system, comprising 84 hospitals across 16 parent organizations with *N* = 25,000 simulated individuals, designed to address the practical question of whether federation should occur at the hospital or organizational level. Across all settings, we benchmark the proposed algorithms against centralized baselines, non-private distributed baselines, encryption-based privacy-enhancing technologies (Froelicher et al. 2021, Geva et al. 2023), and first-order differentially private optimization methods (Rizk et al. 2023), reporting statistical performance, runtime, and privacy-utility tradeoffs as functions of the privacy budget *ε* and the network size *n*.

### 4.1 Differentially Private Distributed Survival Analysis for Clinical Trials

For the first simulation study, we focus on the AIDS Clinical Trials Group (ACTG) Study 175 dataset, with zidovudine (ZDV) as the standard care/control and didanosine (ddI) as the treatment. Throughout this section, we test the treatment effect while adjusting for baseline prognostic covariates (age, CD4 count, Karnofsky performance score, weight, prior antiretroviral exposure, race, and sex).

#### Hypothesis testing

We test whether ddI improves patient survival, that is, 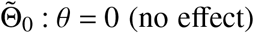against 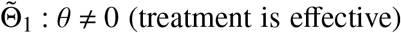, where *θ* is the treatment coefficient of the multivariate proportional hazards model. Because the covariates enter both models identically and cancel in the ratio, the released quantity is a scalar and is exchanged by the belief-propagation algorithms of Section 3 unchanged (see Appendix J for the global sensitivity analysis and other implementation details). We use *n* = 5 fully connected hospitals, a significance level *α* = 0.05, fitting the model with the open-source package <monospace>lifelines </monospace>(Davidson-Pilon 2019) and an *L*_2_ penalty *λ*_reg_|*θ* |^2^ with *λ*_reg_ =0.05. The centralized analysis estimates a ddI log hazard ratio of −0.49 (95% CI [−0.69, −0.28], *P* < 10^−5^; Figure 2B). In Figure 2C and Figure 2D, we observe that the three proposed algorithms (AM, GM, two-threshold) converge to the non-private benchmark as *ε* grows for *ε* ∈ [0.1, 10].

**Figure 2.**
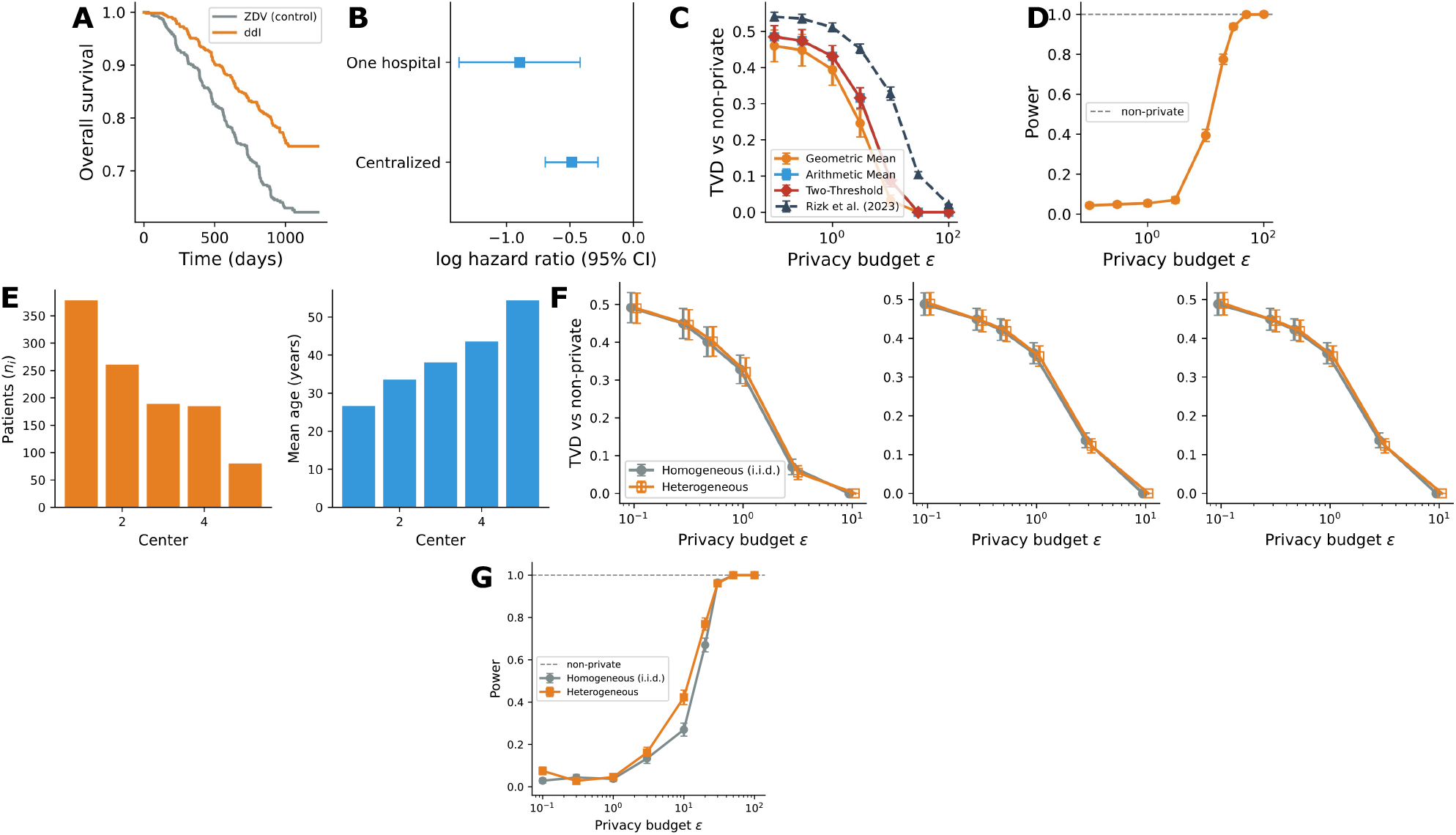
(A-D) Differentially private distributed survival analysis on ACTG 175, testing the ddI treatment effect for *n* = 5 fully connected centers. (A) Kaplan-Meier survival curves for the control (ZDV) and treatment (ddI) arms. (B) ddI log hazard ratio with 95% confidence intervals, fitted centrally and at a single hospital. (C) Total variation distance between the private and non-private beliefs as a function of the privacy budget *ε* for AM, GM, and the two-threshold algorithm, together with the differentially private first-order method of Rizk et al. (2023). (D) Statistical Power of the GM as a function of the privacy budget *ε* at significance level *α* = 0.05. (E-G) Robustness of differentially private distributed survival analysis to federation heterogeneity. (E) A non-i.i.d. partition incorporating empirically-grounded center-size variations and age-shifted case mixes across sites. (F) Total variation distance between the private algorithms (AM, GM, two-threshold) and the non-private algorithm as a function of the privacy budget *ε* under homogeneous vs. heterogeneous partitions. (G) Statistical power of the GM estimator under the homogeneous vs. heterogeneous partitions at a significance level *α* = 0.05. We report 95% confidence intervals taken over 500 repetitions for *T* = 15, *K* = 2.

#### Runtime study

In Appendix J.3, we characterize the computational complexity of our algorithms, running in polynomial time and space. In Appendix J.4, we perform a runtime study on our proposed algorithms and show that our algorithm can run between ∼ 10^−2^ and 10 seconds for values of *n* ranging from *n* = 3 to *n* = 96, which is significantly faster (up to 1000×) than existing methods that rely on homomorphic encryption (Froelicher et al. 2021). Fully homomorphic encryption and multiparty computation are cryptographic techniques that allow computation on encrypted data (by single or multiple servers, respectively) without the need to access decrypted data. The strong cryptographic guarantees of these methods come at a high cost to computational efficiency, and their implementation requires information technology (IT) infrastructure and large-scale, highperformance computational resources across distributed sites, which are beyond the technical capabilities of many organizations.

#### Comparison with first-order methods

First-order distributed optimization methods with differential privacy guarantees offer an alternative approach (Rizk et al. 2023), which, while more general, can be inefficient for distributed maximum likelihood estimation. Figure 2C shows that our algorithms yield answers closer to ground truth compared to the first-order method of Rizk et al. (2023) with significantly smaller errors (details in Appendix J.5).

#### Additional datasets and experiments

In Appendix J.2, we provide experiments with data from clinical trials in patients with advanced cancer, where the task is to determine whether certain biometric indices affect patient survival.

#### Robustness to federation heterogeneity

The adjusted analysis above splits the trial evenly across identical centers over a complete network. Real federated networks are heterogeneous, so we evaluate the method under a highly heterogeneous regime. We assign patients to *n* = 5 centers with unequal sizes drawn from real hospital bed counts (Homeland Infrastructure Foundation-Level Data (HIFLD) 2020) and an age-shifted case mix where we control the mean age of the individuals at each center, maintaining both trial arms at every center (Figure 2E). In Figure 2F and Figure 2G, we observe that the performance of the algorithm remains almost the same under the heterogeneous data split.

### 4.2 Differentially Private Distributed Genetic Association Analysis

Genome-wide association studies exemplify both the necessity and the peril of data sharing. Detecting small per-variant effects demands cohorts larger than any single institution holds, motivating federated consortia. Homer et al. (2008) showed that an individual’s membership in a study can be inferred from published allele frequencies alone, a result that prompted the withdrawal of aggregate GWAS statistics from public access (Zerhouni and Nabel 2008) and still constrains data-sharing policy (Erlich and Narayanan 2014). Nor can leakage be contained by redacting sensitive loci: linkage disequilibrium allows masked genotypes to be reconstructed from neighboring markers, as when James Watson’s withheld APOE status proved recoverable from the adjacent TOMM40 region (Nyholt et al. 2009), ultimately forcing the removal of a 2 Megabase (Mb) window around the locus. In general, syntactic anonymization offers no guarantees against an adversary with side information or knowledge of the genome’s correlation structure. Differential privacy addresses this gap by providing a formal and robust guarantee against arbitrary auxiliary information, which we adopt for federated genetic association analysis in this subsection.

With DP in place, the operative managerial question is no longer whether to protect aggregate statistics, but at what level of institutional aggregation collaboration should occur. Should individual hospitals participate as independent nodes, each contributing their own privatized statistics, or should parent organizations first pool their member hospitals’ data internally and participate as organizational-level nodes? This choice is typically settled through informal negotiation among legal, administrative, and scientific stakeholders, with little quantitative guidance. Our framework brings out the underlying quantitative tradeoffs.

As a case study, we conduct a large-scale regional study to evaluate our method in the context of distributed, differentially private genetic association analysis. Specifically, each simulation replicate consists of *N* = 25,000 individuals genotyped at a panel of 30 independent genomic regions (unlinked loci) of 200kb each, each region spanning roughly fifty variants simulated with the <monospace>msprime </monospace>package (Baumdicker et al. 2022) under a coalescent model that mimics the linkage disequilibrium (LD) structure of European-ancestry samples, the reference population used in large-scale consortia such as the UK Biobank (Sudlow et al. 2015). Six regions are associated and the remaining 24 are null. Each center summarizes every region by its SKAT statistic (Wu et al. 2011), and the task is to detect the associated regions. Following real-world studies on Alzheimer’s disease (He et al. 2021), we consider both rare (Minor Allele Frequency, MAF < 0.01) and common (MAF ≥ 0.01) variants, with a realistic composition of 70% rare and 30% common variants. The six associated regions are assigned a uniform range of per-region heritabilities ranging from 3% to 22%. This mirrors the effect-size architecture of complex traits, whose heritability is carried disproportionately by a few larger-effect loci and spread across many weaker loci, which allows us to show the operating regime of our method.

Next, we construct a realistic network of federating centers from the publicly available US Hospital Locations dataset from Homeland Infrastructure Foundation-Level Data (HIFLD) (2020). We retain all open, acute-care hospitals in New York State lying within the eight counties that comprise the New York City metropolitan area: Manhattan (New York County), Brooklyn (Kings), the Bronx, Queens, Staten Island (Richmond), Nassau, Westchester, and Suffolk. This yields 84 hospitals across 8 counties, totaling 32,989 beds. These hospitals belong to 16 unique organizations (independent hospitals grouped together). We consider two topologies that reflect our main question: *(i)* an organizational-level complete network among the *n* = 16 organizations, assuming that the hospitals within each organization can pool their data together without the need for an agreement, and *(ii)* a hospital-level geographical network among the *n* = 84 hospitals, in which each hospital is connected to the 3-nearest hospitals based on their geographical distance. We operationalize heterogeneity in hospital size by assigning each hospital *i* a simulated cohort of *n*_*i*_ individuals proportional to its total bed count (see Figure 3C). Here, the sensitivity Δ_*i*,Θ_ grows polynomially with the number *M* of SNPs, so we clip each individual’s contribution to bound the sensitivity and add the appropriately calibrated Laplace noise (see Appendices K.2 and K.3).

**Figure 3.**
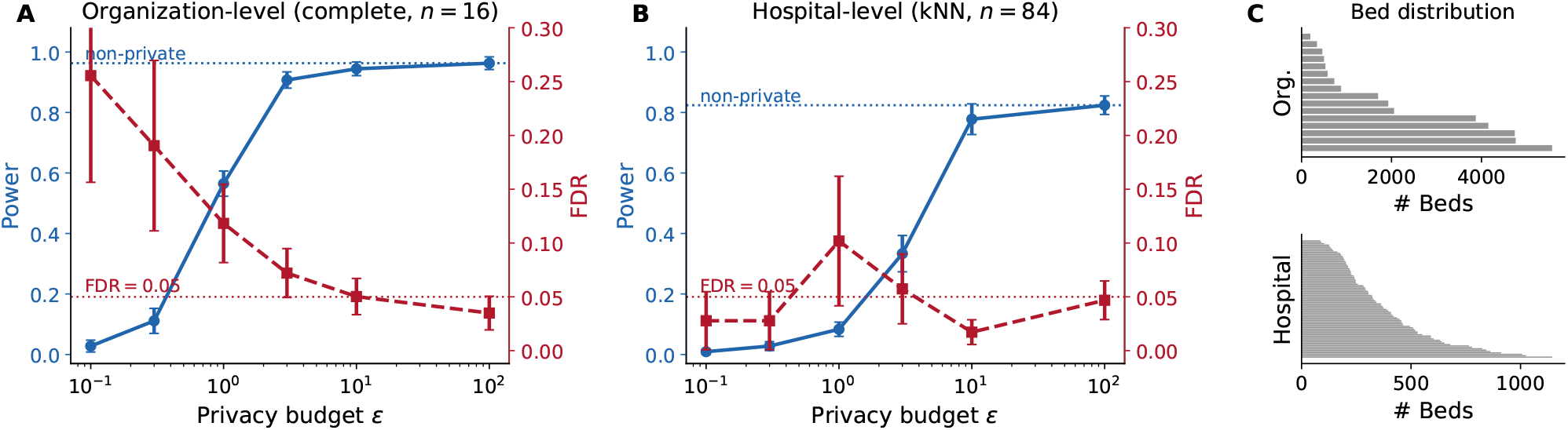
Privacy-utility curves measuring power, FDR (Type I error), for differentially private distributed genetic association analysis under two network architectures derived from the New York City hospital system with *N* = 25, 000 individuals assigned according to hospital sizes shown in Panel (C). (A) Privacy-utility curve of the complete organizational network containing *n* = 16 organizations pertaining to the organizations that the hospitals in the New York City system correspond to. (B) Privacy-utility curve for the geographic hospital network containing *n* = 84 hospitals, where we have assumed that each hospital can exchange information with its 3-nearest hospitals. We observe that in the first case (panel A), the critical value of *ε*, where the DP algorithm has sufficient power (approaching the centralized and the no-DP baselines), and low FDR (less than 5%) is *ε* being between 1 and 10, which is reasonable by widely-documented standards. In the latter case, the hospital network achieves much lower power showing that the organization level federation is better. We have used *T* = 10 and *K* = 2 and report 95% confidence intervals over 3 runs.

Appendix K presents the behavior of the method for different parameters such as the cohort size (*N*), the number of participating centers (*n*), and the network structure, and shows that our method is able to perform inference with a runtime on the order of seconds for a large number of hospitals, which is orders of magnitude smaller than homomorphic encryption methods and with substantially smaller error compared to first order methods.

#### Privacy-utility trade-offs

Figure 3 compares the statistical power and the false discovery rate of the federated DP-GWAS algorithm under two network architectures derived from the New York City hospital system. The organizational-level network achieves statistical power that approaches the centralized baseline, with the false discovery rate controlled below 5% at a critical privacy budget of *ε*^∗^ between 1 and 10. The hospital-level network, by contrast, requires a larger privacy budget and attains a lower power: it federates over many more centers, each of which injects Laplace noise and mixes more slowly over the sparse geographic network. To understand why, it is instructive to examine the privacy-utility curves for both networks as a function of *ε* (Figure 3A and B).

On the one hand, for the organizational-level network, there exists a critical value of *ε* between 1 and 10 at which the algorithm transitions from the strong privacy regime (*ε* < *ε*^∗^), where the region statistics are dominated by noise and statistical power is lower, to a regime where power converges to the non-private baseline (Figure 3A). On the other hand, for the hospital-level network, this critical *ε*^∗^ is much larger, making it less practically deployable (Figure 3B).

When *ε* is below *ε*^∗^, the privatized statistics exchanged between centers are dominated by noise rather than by the true genetic signal, so few associated regions are above the significance threshold and statistical power is low. As *ε* increases above *ε*^∗^, the signal-to-noise ratio improves and power increases toward the non-private centralized baseline at an FDR below the target level.

## 5. Discussion

### Managerial Insights

Our results carry concrete implications for decision-makers at institutions considering participation in distributed collaborative clinical research. Four main themes follow:

#### The critical privacy budget informs decision-making

The privacy budget *ε* can inform decisionmaking: organizations can offer stronger privacy guarantees (smaller *ε*) to hesitant partners at a quantifiable and predictable cost to statistical power and communication rounds, making the privacy-utility trade-off tangible up to a critical level *ε*^∗^. As shown in Figure 3, operating below this critical *ε*^∗^, the algorithm operates at a low signal-to-noise-ratio, yielding low power and high FDR. On the other hand, increasing *ε* above the critical threshold *ε*^∗^ yields high power and low FDR at the expense of privacy. From our experiments, choosing *ε*^∗^ to be approximately between 1 and 10 for the organizational-level network represents a notion of a *minimum deployable privacy budget*.

#### Aggregation strategy and error tolerance

The choice between the AM and GM estimators has a direct interpretation: Organizations in the early-stage screening contexts, where missing a true signal is the costlier error, should prefer the AM estimator, which controls Type-II error and maximizes recall. Those operating in regulatory approval or treatment selection contexts, where false positives carry legal or clinical liability, should prefer the GM estimator, which controls Type-I error and ensures that only well-supported alternatives are selected. Finally, our two-threshold algorithm offers finer simultaneous control over both error types at a modest increase in communication complexity, and is preferable when neither error type is clearly dominant.

#### Implications for network design

The structure of the collaboration network (communication graph) has quantifiable consequences for statistical performance. Denser, more connected networks converge faster, require fewer communication rounds, and achieve a given accuracy at a smaller privacy budget, as reflected in the dependence of our complexity bounds on the second-largest eigenvalue modulus of the adjacency matrix. This reinforces the argument for investing in broader inter-institutional partnerships compared to bilateral agreements. Our runtime comparisons further demonstrate that DP-based collaboration can be deployed with existing computational infrastructure and runs significantly faster than homomorphic encryption methods.

Further, the most actionable insight from our empirical evaluation concerns the unit at which institutions should participate in a federated network. Comparing the organizational-level and hospital-level networks derived from the New York City hospital system reveals significant differences: the organizational network achieves power that approaches the centralized baseline with an FDR below 5% at the critical *ε*^∗^, which is much lower than the hospital-level network.

This divergence reflects three compounding mechanisms: First, individual hospitals hold smaller cohorts, weakening the per-node log-likelihood signal relative to the noise. Second, the larger number of nodes increases communication complexity and introduces more noise accumulation per round. Third, the sparser adjacency matrix of the hospital network slows convergence, requiring more iterations to converge. Thus, the fragmentation of the network into many small nodes degrades privacy-utility performance, and the organizational-level architecture delivers both stronger privacy and better inference by improving the signal-to-noise ratio before any privatized information is shared externally.

Therefore, we recommend a two-tier governance architecture in which hospitals first pool data within their parent organization under existing intra-organizational protocols, typically less burdensome than inter-organizational DUAs and not requiring patient-level re-consent in most jurisdictions, and only organizational nodes participate in the external federated network under formal DP guarantees. This architecture reduces communication complexity, strengthens the formal privacy guarantee offered to patients and regulators, and improves statistical power. In practice, large hospital systems already centrally govern data across their member hospitals and hold IRB approvals covering data use across member institutions, making the organizational node a natural and already existing unit of participation. When deciding how to structure participation, the decision rule is straightforward: if the pooled cohort size of the organizational node is large enough for the signal to dominate the DP noise at *ε* < *ε*^∗^, the organizational-level architecture should be adopted; otherwise, the institution should consider pooling with a peer organization under a bilateral DUA to form larger cohorts, or else admit a larger *ε* (incurring privacy costs) or a reduced statistical power to maintain their current granular node-level participation.

### Limitations and Future Work

Several limitations of the present work merit discussion. First, our theoretical analysis assumes a fixed, known communication network with an irreducible doubly stochastic adjacency matrix; extending the framework to time-varying or randomly failing networks, which are common in practical distributed systems, remains an open problem.

Second, communication complexity scales with the number of rounds *K* and the state space size |Θ|; for very small *ε* or large |Θ|, this can become prohibitive. Developing more communication-efficient aggregation strategies, for example, using the Dirichlet mechanism (Gohari et al. 2021), is an important direction for future work.

Third, our finite-sample guarantees are developed for simple hypotheses, where each state in Θ indexes a single, fully specified likelihood. A natural and important next step is to extend this framework to more complex settings, where each hypothesis is a family of distributions, and the test statistics (e.g., generalized likelihood ratios) must be optimized over that family with appropriate sensitivity bounds for privacy calibration.

Finally, our empirical evaluation focuses on survival and genetic association analyses; validating the framework across a broader range of distributed inference tasks (e.g., differential expression (DE) for biomarker discovery, best-treatment selection on binary or continuous endpoints, and diagnostic classification and disease subtyping) would be promising next steps.

### Concluding Remarks

Motivated by the need to share sensitive clinical data across institutions, we have studied distributed estimation and learning in networks of healthcare centers that collaborate without pooling raw patient records, yet still face privacy risks with respect to both their own data and the beliefs they exchange with their neighbors. We proposed algorithms for distributed MLE and online learning that incorporate rigorous *ε*-DP guarantees through calibrated Laplace noise injection and derived finite-sample bounds on Type-I and Type-II error probabilities that characterize the three-way trade-off among privacy, communication complexity, and inference accuracy. Empirical validation on real-world clinical trial and synthetic genomic data demonstrates that our framework achieves statistical performance that approaches the centralized baseline under practically deployable privacy budgets, at runtimes orders of magnitude faster than cryptographic baselines, and with lower error than existing first-order DP optimization methods. Beyond the technical contributions, our analysis yields actionable guidance for the design of federated research networks, offering a principled quantitative basis for decisions about privacy budgets, aggregation strategies, network topology, and the level at which institutions should participate.

## Data Availability

The data used to support the findings of the paper can be downloaded from the following sources:
* The AIDS Clinical Trials Dataset has been obtained from Kaggle: https://www.kaggle.com/datasets/tanshihjen/aids-clinical-trials (Accessed on February 14, 2025). Original information about the clinical trial can be found at https://clinicaltrials.gov/study/NCT00000625 (Accessed on February 14, 2025).
* The cancer clinical trial dataset has been obtained from Samstein et al. (2019) and is available at https://doi.org/10.1038/s41588-018-0312-8 (Accessed on February 14, 2025).
* The code used to run the simulations can be found at https://github.com/papachristoumarios/dp-social-learning (Accessed on March 10, 2025)

https://doi.org/10.1038/s41588-018-0312-8

https://clinicaltrials.gov/study/NCT00000625

https://www.kaggle.com/datasets/tanshihjen/aids-clinical-trials

## Acknowledgments

M.P. was partially supported by a LinkedIn Ph.D. Fellowship, an Onassis Fellowship (ID: F ZT 056-1/2023-2024), and grants from the A.G. Leventis Foundation and the Gerondelis Foundation. The authors would like to thank the seminar participants at Rutgers Business School, Jalaj Upadhyay, Saeed Sharifi-Malvajerdi, Jon Kleinberg, Kate Donahue, Richa Rastogi, the attendants of the Information Systems Workshop at Arizona State University, Sue Brown, Maytal Saar-Tschechansky, and Vasilis Charisopoulos for their valuable discussions and feedback.

## Appendix

This appendix is organized as follows. First, Appendix A presents a table of notations used throughout the paper, and Appendix B summarizes the communication complexity of our proposed algorithms. Appendix C presents the normative foundations behind the log-linear belief exchange, and Appendix D presents the non-private benchmark algorithm for MLE. Two of the private belief propagation algorithms are deferred to this appendix. We present our two-threshold algorithm for private distributed MLE in Appendix E and our algorithm for differentially private distributed online learning from intermittent streams in Appendix F. The convergence proofs for the private belief exchange algorithms are provided in Appendices G to I. Finally, Appendix J presents the sensitivity analysis and additional experiments supporting our distributed survival analysis for clinical trials, and Appendix K presents those supporting our distributed genetic association analysis.

## Appendix Contents

## Appendix A

**Table of Notations**

Appendix A: Table of Notations
|  |  |
| --- | --- |
| $x$ | scalar |
| $\bar{x}$ | vector |
| $\mathbf{x}$ | element of vector $\bar{x}$ |
| $\ \bar{x}\ _p$ | $\ell_p$-norm of $\bar{x}$ |
| $X$ | matrix |
| $\mathbb{E}[\cdot], \mathbb{V}[\cdot]$ | expectation and variance operators |
| $n$ | number of agents |
| $\varepsilon$ | privacy budget |
| $\alpha, 1 - \beta$ | Type I, Type II error probabilities |
| $\eta$ | $\eta = \max\{\alpha, 1 - \beta\}$ |
| $A$ | communication matrix |
| $\Theta$ | set of states |
| $\Theta^*, \bar{\Theta}$ | set of MLE, non-MLE states |
| $\theta^*, \bar{\theta}, \theta^\circ$ | MLE, non-MLE/false state, true state (in the online learning case) |
| $f^*$ | density of MLE states ($f^* = \vert \Theta^*\vert /\vert \Theta\vert $) |
| $\ell_i(s\vert \hat{\theta})$ | likelihood of agent $i$ given dataset $s$ at state $\hat{\theta}$ |
| $T, K$ | number of iterations, number of rounds |
| $\gamma_{i,t}(\hat{\theta})$ | joint likelihood of agent $i$ given data $s_{i,t}$ of size $n_{i,t}$ at state $\hat{\theta}$ in the non-private regime |
| $\lambda_{i,t}(\hat{\theta}, \check{\theta})$ | joint log likelihood ratio of agent $i$ at iteration $t$ between states $\hat{\theta}$ and $\check{\theta}$ in the non-private regime |
| $\Lambda(\hat{\theta}, \check{\theta})$ | joint log likelihood ratio between states $\hat{\theta}$ and $\check{\theta}$ over all agents and data in the non-private regime |
| $\sigma_{i,t}(\hat{\theta})$ | joint likelihood of agent $i$ given data $s_{i,t}$ of $n_{i,t}$ patients $\hat{\theta}$ in the private regime |
| $\zeta_{i,k,t}(\hat{\theta}, \check{\theta})$ | joint log likelihood ratio of agent $i$ at iteration $t$ at round $k$ between states $\hat{\theta}$ and $\check{\theta}$ in the private regime |
| $Z_k(\hat{\theta}, \check{\theta})$ | joint log likelihood ratio between states $\hat{\theta}$ and $\check{\theta}$ at round $k$ over all agents and data in the private regime |
| $\mu_{i,t}(\hat{\theta})$ | belief of agent $i$ at iteration $t$ on state $\hat{\theta}$ in the non-private regime |
| $\nu_{i,k,t}(\hat{\theta})$ | belief of agent $i$ at iteration $t$ at round $k$ on state $\hat{\theta}$ in the private regime |
| $\phi_{i,t}(\hat{\theta}, \check{\theta})$ | log belief ratio of agent $i$ at iteration $t$ on pair of states $\hat{\theta}$ and $\check{\theta}$ in the non-private regime |
| $\psi_{i,k,t}(\hat{\theta}, \check{\theta})$ | log belief ratio of agent $i$ at iteration $t$ at round $k$ on pair of states $\hat{\theta}$ and $\check{\theta}$ in the private regime |
| $d_{i,t}(\hat{\theta}), d_{i,k}(\hat{\theta})$ | noise variable for agent $i$, state $\hat{\theta}$, and iteration $t$, or round $k$ |
| $\mathcal{D}_{i,t}(\hat{\theta}; \varepsilon), \mathcal{D}_{i,k}(\hat{\theta}; \varepsilon)$ | noise distribution for agent $i$, state $\hat{\theta}$, and iteration $t$, or round $k$ |
| $\delta_{ij}$ | censoring indicator for patient $j$ in center $i$ |
| $\mathbf{x}_{ij}$ | treatment indicator for patient $j$ in center $i$ |
| $t_{ij}$ | survival or censoring time in days for patient $j$ in center $i$ |
| $b_n^* = \vert \lambda_2(A)\vert $ | second-largest eigenvalue modulus of $A$ |
| $a_n^*$ | second largest eigenvalue modulus of $(A + I)/2$, also denoted by $\vert \lambda_2(A + I)\vert /2$ |
| $\Gamma_{n,\Theta}$ | largest likelihood magnitude |
| $\Delta_{i,\Theta}$ | global sensitivity of center $i$ |
| $\Delta_{\Theta}^n$ | maximum of the global sensitivities; $\max_{i \in [n]} \Delta_{i,\Theta}$ |
| $l_{n,\Theta}$ | minimum KL-divergence between non-MLE and MLE states (false and true states in the OL case) |
| $V_{n,\varepsilon,\Theta}$ | sum of standard deviations of the noises over all agents |
| $Q_{n,\Theta}$ | maximum standard deviation of KL-divergence for any agent and any pair of states |

## Appendix B

**Summary Table for the Communication Complexity of MLE Algorithms**

**Table B.1.** Communication complexity of distributed inference algorithms for a fixed privacy budget *ε* > 0 and target maximum Type I/Type II error *η* ∈ (0, 1). We use O_*a*1,…,*aN*_ (·) to denote the big-O notation where the constants are allowed to depend on *a*_1_, …, *a*_*N*_ . The overhead of introducing privacy is denoted in blue. For compactness in notation, *η* refers to max{*α*, 1 − *β*} and *ϱ* refers to min{*ϱ*^AM^, *ϱ*^GM^} (see Theorem 2 for closed-form lower bounds with explicit dependence on the AM and GM thresholds *ϱ*^AM^ and *ϱ*^GM^ and error probabilities *α* and 1 − *β*). In the case of the two-threshold algorithm, *π* refers to min{*π*_1_, *π*_2_}, and *q* refers to min{*q*_1_, 1 − *q*_2_} (see Theorem 4 and Algorithm 3 for details of how the two thresholds are determined in terms of parameters *π*_1_, *π*_2_ and *q*_1_, *q*_2_). The AM/GM algorithms have matching communication complexities with the two-threshold algorithm whenever *q* = O(1) and 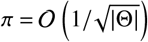. The non-DP results are due to Rahimian and Jadbabaie (2016).

| Setting | With DP (ours) | Without DP (Rahimian and Jadbabaie 2016) |
| --- | --- | --- |
| Communication Complexity with respect to $\varepsilon, \eta$ | | |
| AM/GM | $O_{\vert \Theta\vert ,n}(\log(1/\eta)(\log(1/\eta) + \log(1/\varepsilon) + \log(1/\varrho)))$ | $O_{\vert \Theta\vert ,n}(1)$ |
| Two-Threshold | $O_{\vert \Theta\vert ,n}\left(\frac{\log(1/\eta)}{\pi^2}(\log(1/\eta) + \log(1/\varepsilon) + \log(1/q))\right)$ | $O_{\vert \Theta\vert ,n}(1)$ |
| Communication Complexity with respect to $n$ | | |
| AM/GM | $O_{\vert \Theta\vert ,\varepsilon,\eta}\left(\max\left\{\log\left(\frac{n}{I_{n,\Theta}}\right), \frac{\log\left(\frac{n(\Gamma_{n,\Theta} + \Delta_{\Theta}^n)}{\varrho}\right)}{\log(1/a_n^*)}\right\}\right)$ | $O_{\vert \Theta\vert }\left(\max\left\{\log\left(\frac{n}{I_{n,\Theta}}\right), \frac{\log(n\Gamma_{n,\Theta})}{\log(1/a_n^*)}\right\}\right)$ |
| Two-Threshold | $O_{\vert \Theta\vert ,\varepsilon,\eta}\left(\frac{1}{\pi^2} \max\left\{\log\left(\frac{n}{I_{n,\Theta}}\right), \frac{\log\left(\frac{n(\Gamma_{n,\Theta} + \Delta_{\Theta}^n)}{q}\right)}{\log(1/a_n^*)}\right\}\right)$ | $O_{\vert \Theta\vert }\left(\max\left\{\log\left(\frac{n}{I_{n,\Theta}}\right), \frac{\log(n\Gamma_{n,\Theta})}{\log(1/a_n^*)}\right\}\right)$ |
| Communication Complexity with respect to $\vert \Theta\vert $ | | |
| AM/GM | $O_{n,\varepsilon,\eta}\left(\vert \Theta\vert \log \vert \Theta\vert \left(\log \vert \Theta\vert + \log\left(\frac{\Gamma_{n,\Theta} + \vert \Theta\vert \log \vert \Theta\vert \Delta_{\Theta}^n}{\varrho}\right)\right)\right)$ | $O_n(\log \Gamma_{n,\Theta})$ |
| Two-Threshold | $O_{n,\varepsilon,\eta}\left(\frac{\log \vert \Theta\vert }{\pi^2} \left(\log \vert \Theta\vert + \log\left(\frac{\Gamma_{n,\Theta} + \vert \Theta\vert \log \vert \Theta\vert \Delta_{\Theta}^n}{q}\right)\right)\right)$ | $O_n(\log \Gamma_{n,\Theta})$ |

## Appendix C

**Differentially Private Log-Linear Belief Updates and Opinion Pools**

The fastest way to (asymptotically) compute the joint log-likelihood would be for the agents to run a linear consensus algorithm (DeGroot 1974, Olshevsky 2017) in the (privatized) log-likelihood space for each *θ* ∈ Θ. This rule is equivalent to performing log-linear updates (i.e., geometric means) in a (non-Bayesian) belief space (Rahimian and Jadbabaie 2016). On the other hand, simple DeGroot (linear) averaging of beliefs converges to the average of initial likelihoods (rather than log-likelihoods) and can give biased estimates if used for maximum likelihood estimation. Kayaalp et al. (2024) and Lalitha et al. (2018, Corollary 2) point out that log-linear learning converges faster than linear learning in the belief space. Marden and Shamma (2012) provide guarantees on the percentage of time that the action profile will be a potential maximizer in potential games using log-linear best-response dynamics.

The log-linear updates that we use to exchange belief statistics among neighboring agents also have parallels as opinion pools to combine probability distributions from multiple experts in the decision and risk analysis literature (Clemen and Winkler 1999). Abbas (2009), in particular, proposes scoring rules based on the KL divergence between the expert distributions and aggregate beliefs that yield linear and loglinear aggregates as the solution to the expert aggregation problem: If KL divergence is computed from the aggregate belief to the expert beliefs then we obtain log-linear updates, otherwise (i.e., if the KL divergence is computed from the expert beliefs to the aggregate belief) we obtain linear updates. Denoting expert beliefs by ***v*** _*j*_ with weights *a*_*i j*_, *j* ≠ *i*, the log-linear belief aggregate *v*^★^ sets the belief in every state ***v***(*θ*) proportionally to 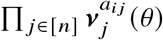and is the solution to arg min ∑_*j*≠*i*_ *a*_*i j*_ *D*_*KL*_ ***v***_*i*_ |***v*** _*j*_. Such rules are especially sensitive to experts who put zero or small beliefs on some states and can be useful for rejecting non-MLE states or null hypotheses. In this section, we use the framework of Abbas (2009) with explicitly modeled privacy costs to justify our update rules beyond the algorithmic performance framework of Section 2.2 and our theoretical guarantees in Section 3.

Following Abbas (2009), consider a decision maker (*i*) who is interested in choosing the best option among a set of alternatives (Θ), given access to their own private information (***s***_*i*_) and other opinions from *n* experts. The decision maker forms a time-varying public belief ***v***_*i,t*_ ∈ Simplex(Θ) over Θ for treatments that capture all expert distributions and its own data. Initially (at *t* = 0), the agent forms an intrinsic belief ***γ***_*i*_ ∈ Simplex(Θ) based on its own data, i.e., 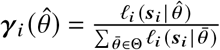for all 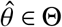. In subsequent steps (*t* ≥ 1), the decision maker chooses an expert at random with probability *a*_*i j*_ corresponding to the strength of the connection in the communication matrix *A* and pays *C*_*i*_ to consult with them and *C*_*i*_ to consult its own data and incurs a revenue *R*_*i*_ regardless of its decision. Since the decision maker consults its own data and communicates its beliefs with others, it must protect its own datasets due to privacy risks.

The *ε*-DP mechanism ℳ_*i*_, satisfying Definition 1, applied to ***γ***_*i*_ produces ℳ_*i*_ (***γ***_*i*_) which is the privatized belief over the state space |Θ|. Putting everything together, the expected payoff of agent *i* at time *t* is given as (see Eq. (9) in Abbas (2009)):

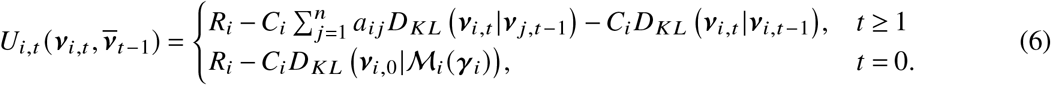

To implement the mechanism ℳ_*i*_, the decision maker draws appropriately chosen, without loss of generality, zero-mean noise variables 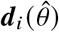for each state 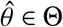and adds them to the corresponding loglikelihoods log 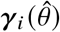. Thus, ℳ_*i*_ (***γ***_*i*_) can be written as the product of the two distributions ***γ***_*i*_ and *D*_*i*_, where 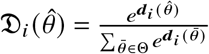. If we write ***v*** as the product of ***v*** and the uniform distribution *U*_Θ_ on Θ, then we can use the properties of the KL divergence to write *D*_*KL*_ ***v***_*i*,0_|ℳ_*i*_ (***γ***_*i*_) = *D*_*KL*_ ***v***_*i*,0_ |***γ***_*i*_ + *D*_*KL*_ (*U*_Θ_|*D*_*i*_). The term *D*_*KL*_ (*U*_Θ_|*D*_*i*_) corresponds to th e privacy loss for the utility in Equation (6) and equals

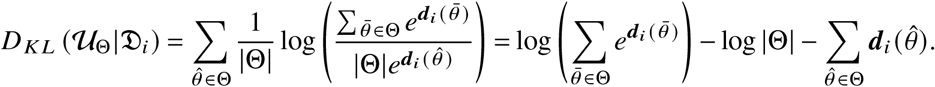

Taking the expectation over M*i* we get 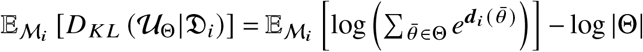. Since the KL divergence is nonnegative, the expected privacy loss with respect to ℳ_*i*_ is minimized whenever the noise variables 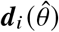are independent of each other and their distribution does not depend on 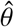, i.e.,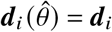.

To satisfy the *ε*-DP requirement in Definition 1, the noise ***d***_*i*_ is set according to the Laplace mechanism with scale 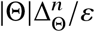and the per-state budget allocation detailed in Section 2.2.

In Appendix J.1, we bound the sensitivity (5) for the proportional hazards model. More generally, the dataset structure may be such that the sensitivity is unbounded when computed globally over the dataset space. In such cases, an additive relaxation of the privacy constraints in Definition 1 can be used with a smoothed notion that looks at sensitivity locally at the realized dataset values (Nissim et al. 2007). The Laplace mechanism possesses further benefits since it minimizes the convergence time of the belief exchange algorithms (cf. Section 3 and prior results by Papachristou and Rahimian (2025) and Koufogiannis et al. (2015)). The best update rule that optimizes the time-varying utility of Equation (6) yields the log-linear update rules (Abbas 2009, Proposition 4).

The above can be extended to the online setting where data ***s***_*i,t*_ with *n*_*i,t*_ ≥ 0 data points for each agent *i* are intermittently arriving at each iteration *t*, by introducing a time-varying utility and measuring the likelihood at each iteration *t*. In that regime, first, the agents form the probability measure ***γ***_*i,t*_ such that 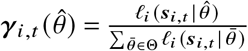for all 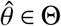(when *n*_*i,t*_ = 0 we set 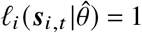for all 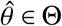). Then the agents have the time-varying utility:

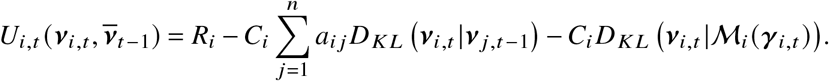

Optimizing this utility yields log-linear learning, which we leverage in our OL Algorithm 5.

## Appendix D

**Non-private Benchmark for Distributed MLE**

In the non-private regime, agents form beliefs 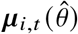over the states 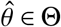at every iteration *t* and exchange their beliefs with their neighbors until they can detect the MLE. Rahimian and Jadbabaie (2016) give Algorithm 2 for belief exchange and prove that beliefs converge to a uniform distribution over the MLE set (see Theorem 1 in their paper).

### Algorithm 2

Non-Private Distributed MLE

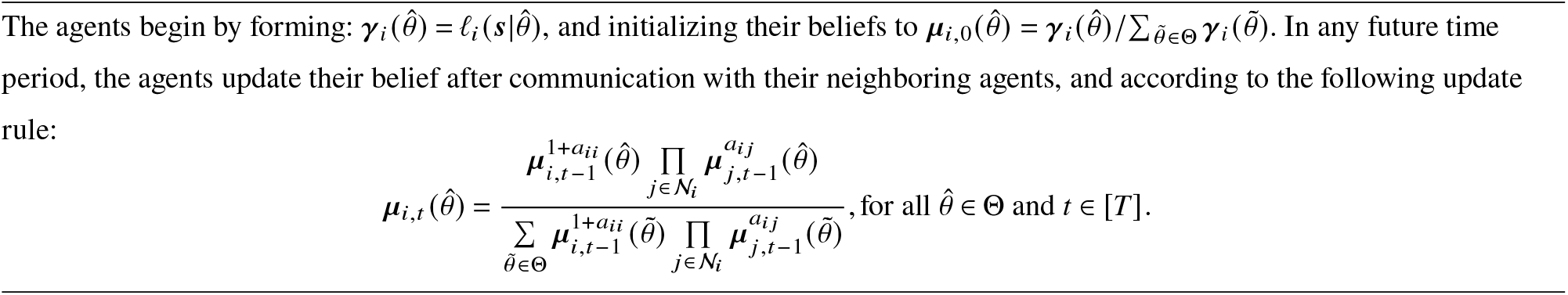

The convergence of this algorithm relies on studying the behavior of the log-belief ratio between two states 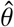and 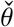, whose dynamics are governed by the powers of the primitive matrix *A* + *I*, see (Rahimian and Jadbabaie 2016, Theorem 1), with modulus-ordered eigenvalues 0 < |*λ*_*n*_ (*A* + *I*) | ≤ · · · ≤ |*λ*_2_(*A* + *I*) | < *λ*_1_(*A* + *I*) = 2. Specifically, Rahimian and Jadbabaie (2016, Theorem 1) state that there is a dominant term due to the maximum eigenvalue *λ*_1_(*A* + *I*) = 2 that dominates the log-belief ratios for large *t*, and *n* − 1 terms that decay exponentially with rate (|*λ*_2_(*A* + *I*) |/2)^*t*^ . The dominant term depends on the difference of the log-likelihoods between the states 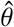and 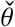, namely 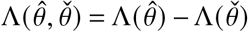. Subsequently, the log-belief ratio approaches −∞ as *t* → ∞ whenever 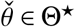, and we can recover the MLEs.

## Appendix E

**The Two-Threshold Algorithm for Private Distributed MLE**

In Algorithm 3, we provide a two threshold algorithm to simultaneously control both Type I and Type II error probabilities, with more design flexibility that comes at an increased cost of communication complexity. The analysis of Theorem 1 shows that in a given round *k*, the probability that an MLE state *θ*^★^ ∈ Θ ends up positive as *T* → ∞ is at least 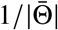, so in expectation at least 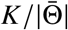rounds will yield a positive belief. On the other hand, we know that for a non-MLE state 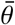, in expectation, at most (1 − 1/|Θ^★^|)*K* trials will come up heads. For brevity, we define *p*_1_ = 1 − 1/|Θ^★^| and 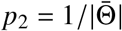Moreover, we define 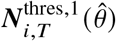(resp.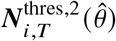) as the (average) number of times the belief 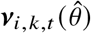exceeds a threshold 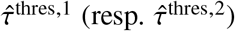for *k* ∈ [*K*].

Because 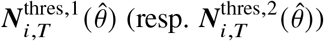is an average of independent indicator variables for all 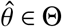, the Chernoff bound indicates that it concentrates around its mean 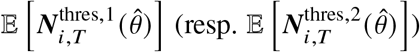. This prompts the development of the following simple algorithm, which resembles boosting algorithms such as AdaBoost (Freund and Schapire 1997). The algorithm uses two thresholds *τ*^thres,1^ and *τ*^thres,2^ to estimate the MLE states as those whose beliefs exceed 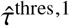and 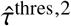at least *τ*^thres,1^ and *τ*^thres,2^ times, leading to output sets 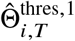and 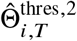, respectively.

We first present an asymptotic result (proved in Appendix G.4):

### Theorem 3.

*Consider Algorithm 3 run with state-independent noise distributions* 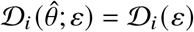*that satisfy ε-DP, and set p*_1_ = 1 − 1/|Θ^★^| *and* 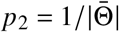. *Then, for every center i* ∈ [*n*], *as ϱ*^thres,1^ → ∞ *and ϱ*^thres,2^ → ∞:

- *for any π*_1_ > 0, *with τ*^thres,1^ = (1 + *π*_1_) *p*_1_ *and* 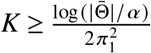, *we have* 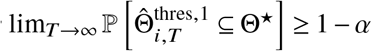;
- *for any π*_2_ >0, *with τ*^thres, 2^ = (1− *π*_2_) *p*_2_ *and* 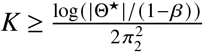, *we have* 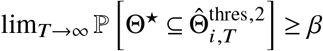.

Similarly to Theorem 1, the estimator 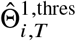has a low Type I error, and the estimator 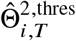has a low Type II error. If agents do not know |Θ^★^|, as long as they choose thresholds 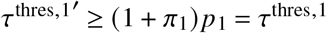and 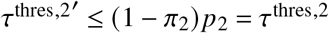, they can obtain the same guarantees. For example, one can choose the following upper and lower bounds to set the thresholds: 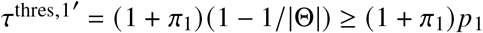and 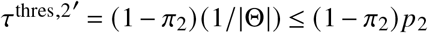. Also, running the algorithm for 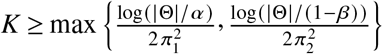we can guarantee that as *T* → ∞, we have 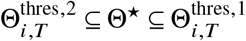with probability at least *β* − *α*

Since the counts for MLE and non-MLE states concentrate around means separated by *p*_2_ − *p*_1_, perfect detection of Θ^★^ is achievable when the two modes are sufficiently separated, i.e., *p*_2_ > *p*_1_, as the following corollary shows:

### Corollary 1

(Exact Recovery with a Single Threshold)

*If the density of MLE states f* ^★^ *satisfies:*

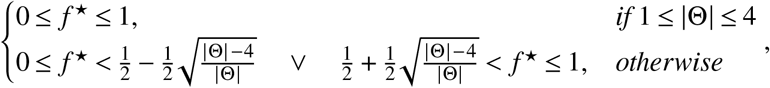

*then setting the thresholds equal, τ*^thres,2^ = *τ*^thres,1^ *and* 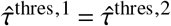*(so that* 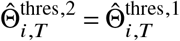*), with* 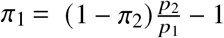*for some π*_2_ > 0 *and* 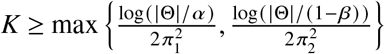, *yields* 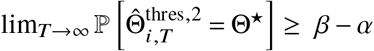*for every center i* ∈ [*n*].

By performing an analysis similar to Theorem 2, we derive a non-asymptotic result (proved in Appendix G.5):

### Theorem 4.

*Let q*_1_, *q*_2_ ∈ (0, 1) *and ϱ*^thres,1^, *ϱ*^thres,2^ > 0. *Then, for Algorithm 3, the following hold for every center i* ∈ [*n*]:

- *For any π*_1_ > 0, if *τ* ^thres,1^ = (1 + *π*_1_) *q*_1_, 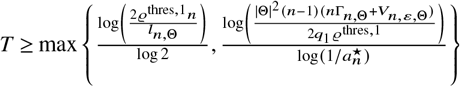, *and* 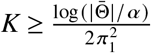*then* 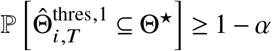.
- *For any π*_2_ > 0, if *τ* ^thres,2^ = (1 + *π*_2_) *q*_2_, 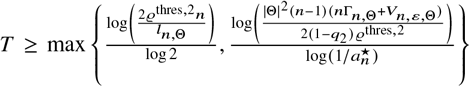, *and* 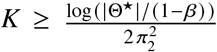, *then* 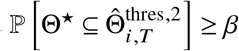.

### Algorithm 3

Private Distributed MLE (Two Threshold Algorithm)

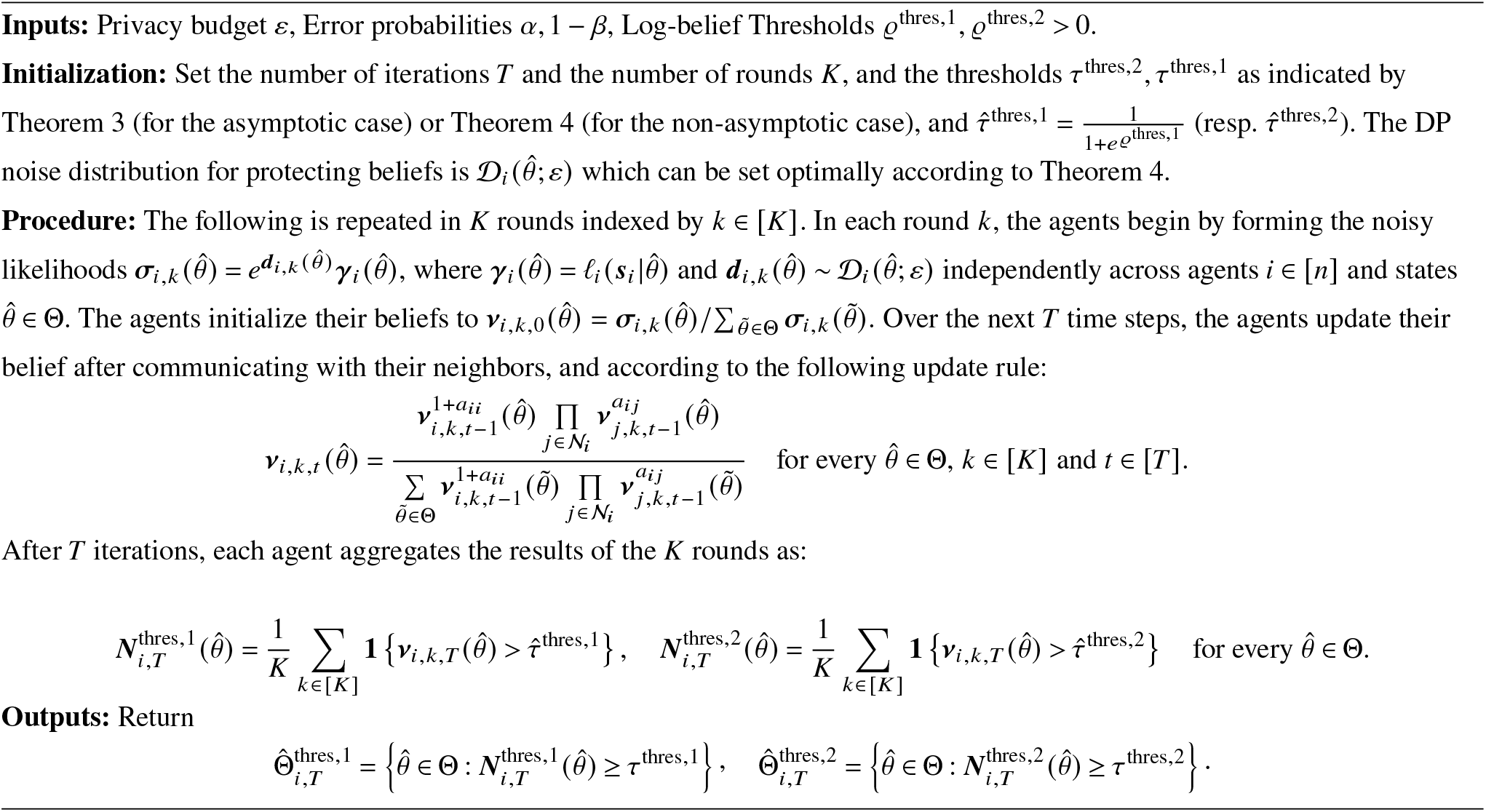

- *The noise distributions that minimize the convergence time T for both estimators are the Laplace distributions* 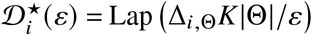.

To minimize the number of iterations, we can pick the thresholds as

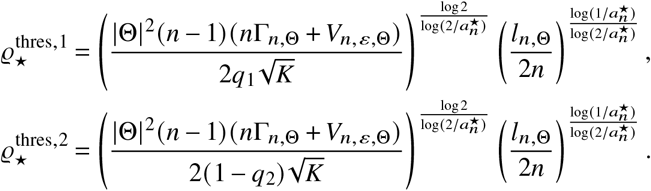

As in Corollary 1, perfect recovery is also achievable with a single threshold when *q*_2_ > *q*_1_ (setting *π*_1_ = (1 − *π*_2_)*q*_2_/*q*_1_ − 1); communication complexity is then minimized by taking 1 − *q*_2_ as close as possible to *q*_1_, but exact recovery costs a polylogarithmic factor in 1/|1 − *q*_2_ − *q*_1_|.

## Appendix F

**Extension to Private Distributed Online Learning from Intermittent Streams**

In the online setting, we consider a network of centers making streams of observations intermittently over time. At each time step *t*, center *i* observes ***s***_*i,t*_ that are distributed according to a model 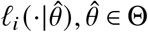. In the online setting, we assume that the data are generated according to a true parameter *θ*^°^ ∈ Θ, which is uniquely identifiable from the collective observations of the centers. The collective identifiability condition can be expressed as 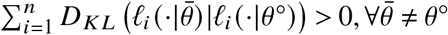. Note that, unlike the MLE setting, where centers can increase the power of their inferences by exchanging belief statistics, in the online setting, each center alone has access to an infinite signal stream.

An informational advantage of collective inference in the online setting is apparent where centers individually face identification problems; for example, for a center *i* ∈ [*n*], a pair of states (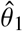and 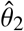) may induce the same distribution of observations, making them indistinguishable from the perspective of center *i*. In such cases, by exchanging beliefs, centers can benefit from each other’s observational abilities to collectively identify the truth even though they may face an identification problem individually.

### The Non-private Online Learning from Benchmark

Rahimian and Jadbabaie (2016) give Algorithm 4 for the agents to determine the true state *θ*^°^ from their streams of observations in the online learning regime, and prove that this algorithm converges to learning the true state asymptotically. Their argument relies on the fact that the time average of the log-belief ratio between any state 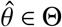and the true state *θ*^°^, 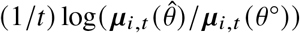, converges to the weighted sum of the KL divergences of all agents, which is less than zero if the models are statistically identifiable.

#### Algorithm 4

Non-Private Distributed Online Learning

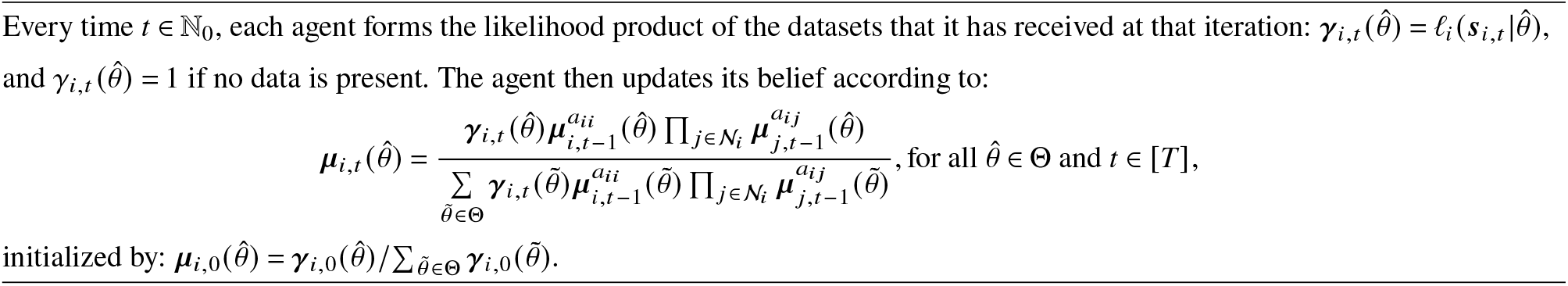

Algorithm 5 facilitates differentially private distributed learning in an online setting, where centers receive datasets at each time step *t*, calculate the likelihood 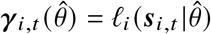and its noisy version 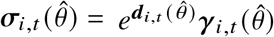to preserve DP, and then aggregate it with their self- and neighboring beliefs from iteration *t* ™ 1. Unlike the MLE setup, for online learning, we do not need to repeat the algorithm in multiple rounds because DP noise cancels out as *T* → ∞.

#### Algorithm 5

Private Distributed Online Learning

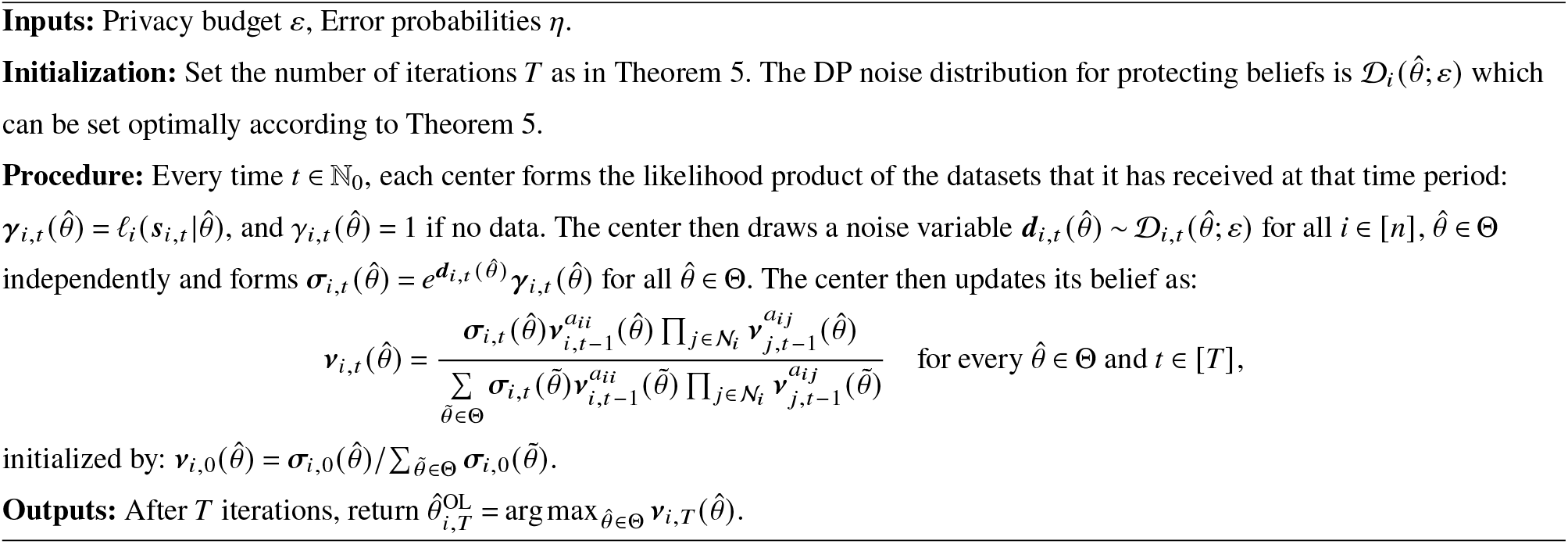

In addition to the global sensitivity parameters Δ_*i*,Θ_ and 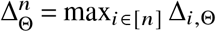(which determine the privacy noise level), the finite-time convergence and implied communication complexity bounds for Algorithm 5 depend on the following statistical properties of the information environment (similarly to Equations (3) and (4), which we encountered in the main text for MLE):

- The minimum divergence between the true state *θ*^°^ and any other state 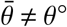:

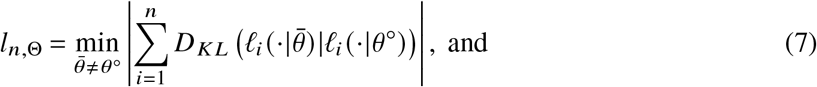
- The maximum standard deviation of the log-likelihood ratio statistics:

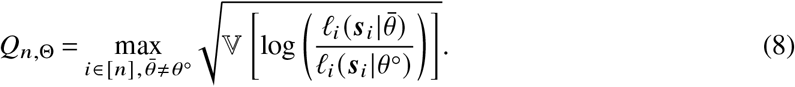

The asymptotic dynamics of beliefs under Algorithm 5 become clear by noticing that the log-belief ratio between *θ*^°^ and any other state 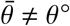converges to 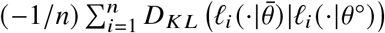, and the effect of DP noise disappears as a consequence of the Césaro mean and the weak law of large numbers. Therefore, centers learn the true state *θ*^°^ exponentially fast, asymptotically (one can show that the asymptotic exponential rate is −*l*_*n*,Θ_/*n*, where *l*_*n*,Θ_ is given by Equation (3)).

The next theorem characterizes the non-asymptotic behavior of Algorithm 5 (proved in Appendix I):

#### Theorem 5.

*Suppose the collective identifiability condition* 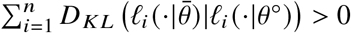*holds for all* 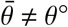. *Let l*_*n*,Θ_ *and Q*_*n*,Θ_ *be the statistical properties of the datasets defined in Equations* (7) *and* (8), *and denote the second-largest eigenvalue modulus (SLEM) of A* 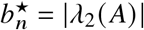. *Then running Algorithm 5 for* 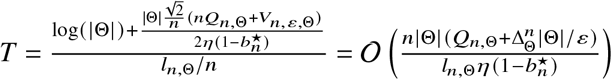*iterations yields* 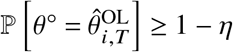*for every center i* ∈ [*n*]. *Moreover, the noise distributions that minimize the runtime are the Laplace distributions* 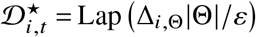, *and the resulting estimates are ε-DP with respect to the private datasets*.

Note that similarly to distributed MLE algorithms, here we also need to scale the privacy budget by |Θ| to account for the fact that the algorithm accesses the private datasets and adds noise for each of the |Θ| states, providing an attacker with as many observations of the private datasets.

The following table compares the communication complexity of distributed online learning for a fixed privacy budget *ε* > 0 in Theorem 5 against the non-private benchmark due to Rahimian and Jadbabaie (2016):

**Table F.1.**
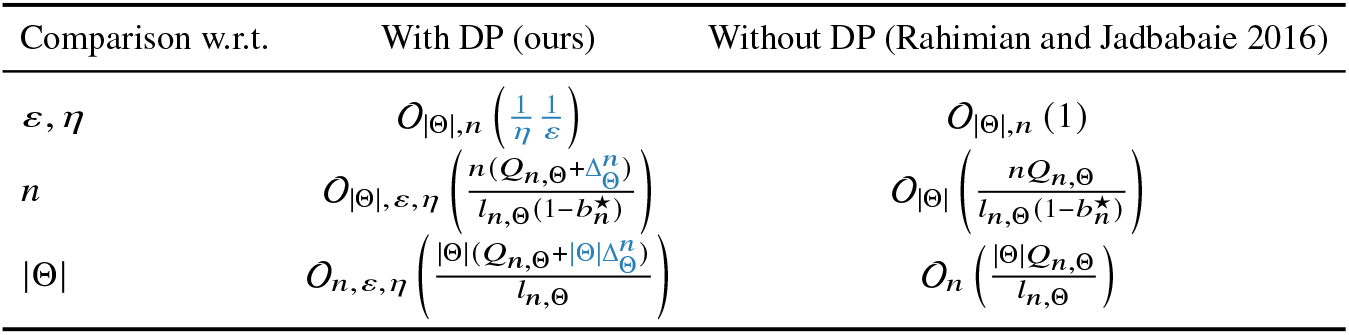
Communication complexity of private and non-private distributed online learning. The overhead of introducing privacy is outlined in blue.

## Appendix G

**Proofs and Convergence Analysis for Private Distributed MLE**

### G.1. Log-Belief Ratio Notations

Let 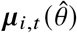 be the beliefs for the nonprivate system, and let 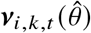 be the private estimates for agent *i* ∈ [*n*], at the time step *t* ≥ 0 of round *k* ∈ [*K*]. For the n on-private algorithm (Algorithm 2) all pairs 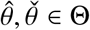 we let 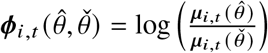 and 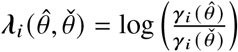 be the log belief ratios and the log-likelihood ratios of the agent *i* ∈ [*n*] in the time step *t* ≥ 0. For the private algorithm (Algorithm 1) we let 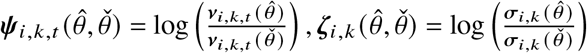, and 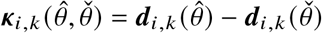 be the private log-belief ratio, log-likelihood ratio and log of the noise ratio between states 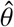 and 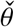 for agent *i* ∈ [*n*] at time step *t* ≥ 0 of round *k* ∈ [*K*], and denote their vectorized versions by 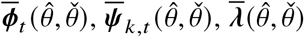, and 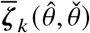, respectively, where the vectorization is over the agents *i* ∈ [*n*]. We define 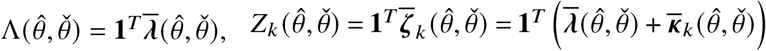.

We say that a stochastic process {*X*_*t*_}_*t* ∈ℕ_ converges in *L*_2_ to *X*, and write 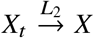, if and only if lim_*t*→∞_ E [∥*X*_*t*_ − *X* ∥_2_] = 0. Convergence in *L*_2_ implies convergence in probability (i.e.,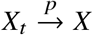) due to Markov’s inequality.

To prove the results, we need the following auxiliary lemma:

#### Lemma 1.

*Let X*_1_, …, *X*_*n*_ *be i*.*i*.*d. draws from a distribution* D. *Then, for every i* ∈ [*n*], *we have* ℙ[*X*_*i*_ ≥ max_*j*≠*i*_ *X* _*j*_] = 1/*n*.

Let *E*_*i*_ = {*X*_*i*_ ≥ max_*j*≠*i*_ *X* _*j*_ }. Note that because *X*_1_, …, *X*_*n*_ are i.i.d. ℙ[*E*_1_] = ℙ[*E*_2_] = · · · = ℙ[*E*_*n*_]. More-over, note that since the maximum is unique, we have that *E*_1_, …, *E*_*n*_ are a partition of the sample space. Therefore, we get ∑_*i*∈[*n*]_ ℙ[*E*_*i*_] = 1, and subsequently ℙ[*E*_*i*_] = 1/*n*. Q.E.D.

### G.2. Proof of Theorem 1: Asymptotic Behavior of the AM/GM Algorithm

Similarly to (Rahimian and Jadbabaie 2016, Theorem 1), we have that for any *k* ∈ [*K*]

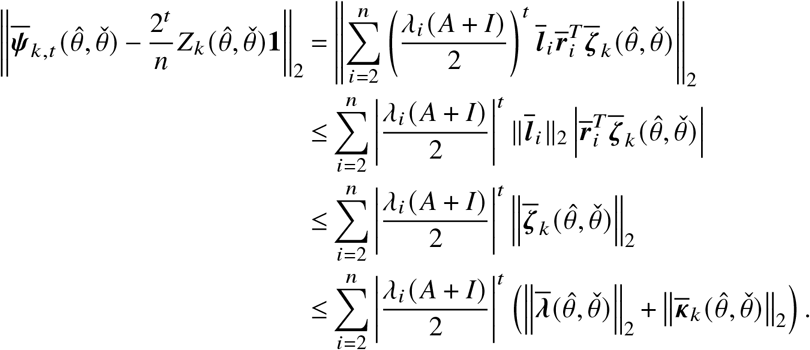

Taking expectations and applying Jensen’s inequality, we can bound the above as

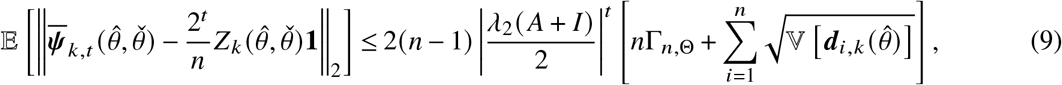

where 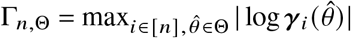 and |*λ*_2_(*A* + *I*) |/2 < 1 is the SLEM of (*A* + *I*)/2, henceforth also denoted by 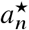, *A* is irreducible doubly stochastic, so *A* + *I* is primitive and 0 < |*λ*_*n*_ (*A* + *I*) | ≤ |*λ*_*n*−1_(*A* + *I*) | ≤ · · · ≤ |*λ*_2_(*A* + *I*) | < *λ*_1_(*A* + *I*) = 2. Note that the right-hand side in Equation (9) goes to 0 as *t* → ∞, which implies that (by Markov’s inequality) 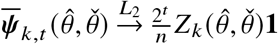.

Let *θ*^★^ ∈ Θ^★^ and 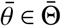. Based on our definition of Λ(·, ·) we should have 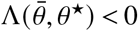 and the corresponding non-private algorithm would have 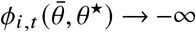 for every 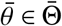 and *θ*^★^ ∈ Θ^★^. However, when noise is introduced, there could be mistakes introduced in the log-belief ratios, even if 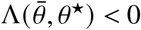 we can have 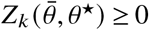 which implies 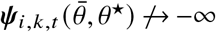.

#### AM estimator

For all 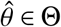 we let 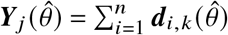. We focus on a single *θ*^★^ ∈ Θ^★^. It is easy to show that because the noise is independent across the agents, and i.i.d. between the states for a given agent *i*, then 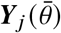 and ***Y***_*j*_ (*θ*^★^) are also i.i.d. for all 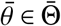, and the refore we have that (see Lemma 1)

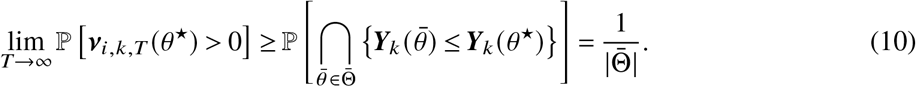

Therefore, for the AM estimator, we have the following:

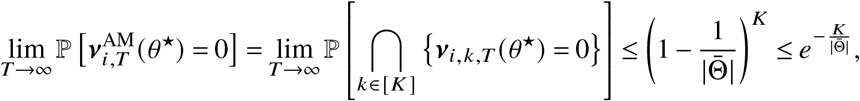

and the failure probability of the AM estimator can be calculated by applying the union bound as

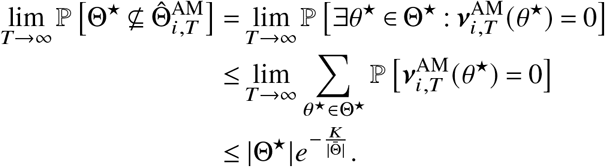

Setting 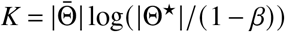, we can ensure that the Type II error rate is at most 1 − *β*.

#### GM estimator

Similarly, for 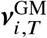 we have the following:

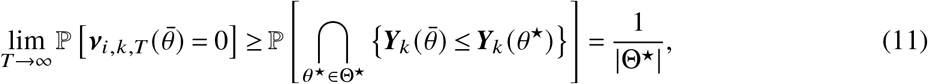

for all 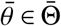 and subsequently

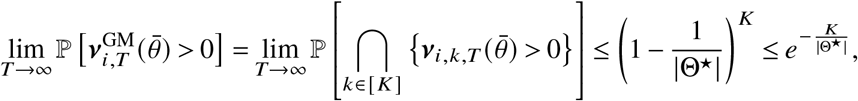

and

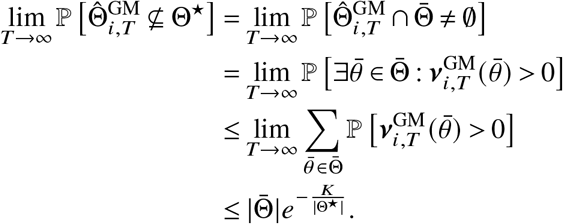

Therefore, selecting 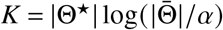 produces a Type I error rate of at most *α*.

#### Negative Impact of *K* on AM Estimator

From the above we know that setting 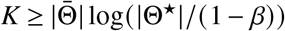 we have that 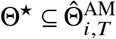 with probability at least *β* as *T* → ∞. We call this “good event” *E*^AM^. Conditioned on *E*^AM^, we have that:

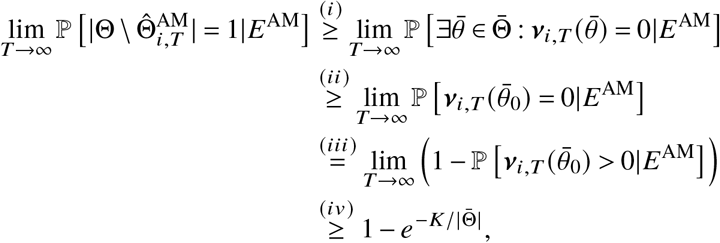

where we have used the following facts: (i) conditioned on *E* a rejected state can only belong to 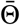, (ii) the probability of the union of events over 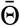 is a superset of the event 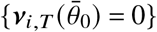 for any fixed 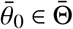, (iii) complement of the event, and (iv) using Equation (10). Therefore

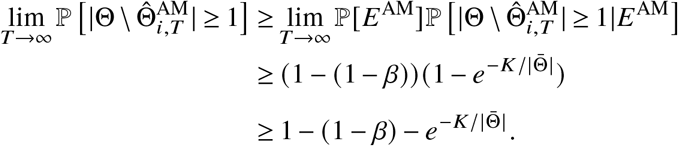

#### Negative Impact of *K* on GM Estimator

From the above we know that setting 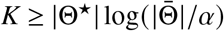 we have that 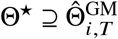 with probability at least 1 − *α* as *T* → ∞. We call this “good event” *E*^GM^. Fix some 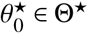. Conditioned on *E*^GM^, we have that:

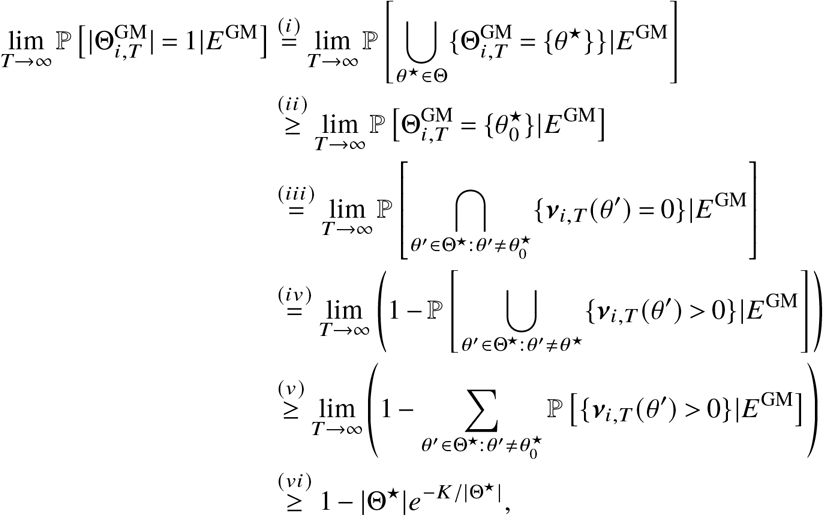

where we have used the following facts: (i) definitiona of the 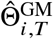 being a signleton conditioned on *E*^GM^, (ii) the fact that the union of events is a superset of a single event, (iii) the fact that the resulting state is a singleton if and only if all other states from Θ^★^ are rejected, (iv) complement of the event, (v) union bound over Θ^★^ \ {*θ*^★^}, and (vi) Equation (11). Finally, combining everything we get that:

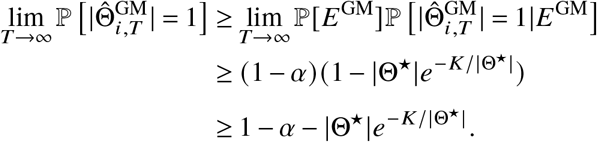

#### Differentially Private Outputs

Due to the postprocessing property of DP (see Proposition 2.1 of Dwork and Roth (2014)), the resulting estimates are *ε*-DP with respect to the private datasets. Q.E.D.

### G.3. Proof of Theorem 2: Finite-Time Guarantees for AM/GM Algorithm

For simplicity of exposition, we have set *ϱ*^AM^ = *ϱ*^GM^ = *ϱ* which corresponds to *τ*^AM^ = *τ*^GM^ = *τ*.

#### GM estimator

We let 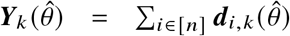. We define the event 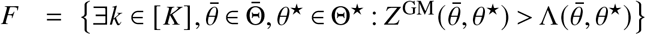. We have that

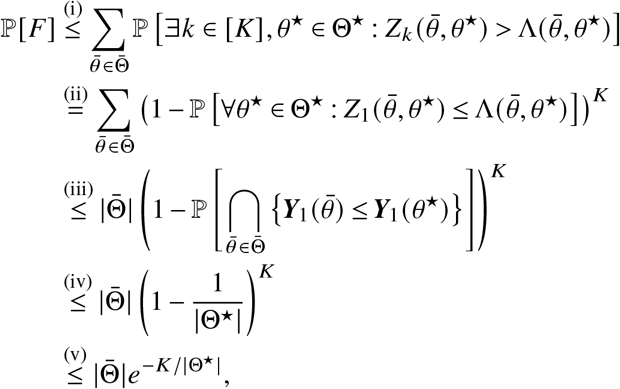

where the above follows (i) from the application of the union bound, (ii) using the fact that the *K* rounds are independent, (iii) the fact that 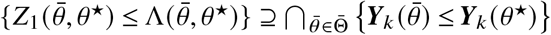 and that 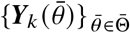 and 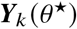 are i.i.d., (iv) Lemma 1, and (v) 1 + *x* ≤ *e*^*x*^ for all *x* ∈ ℝ.

For each round *k* of the algorithm and each pair of states 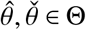, the following holds for the log-belief ratio of the geometrically averaged estimates:

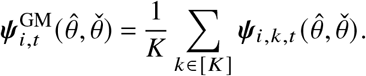

We let 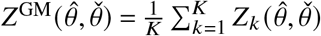. By the triangle inequality and Theorem 1

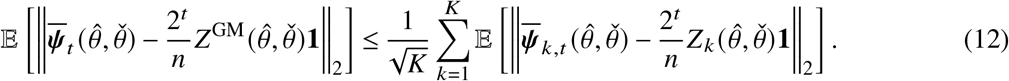

For every round *k* ∈ [*K*],

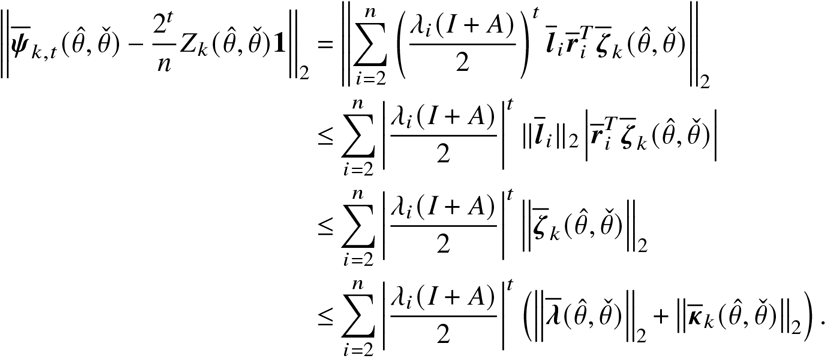

Taking expectations and applying Jensen’s inequality, we can bound the above as

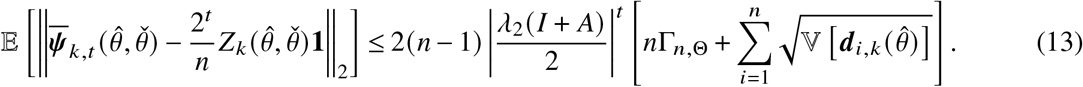

Using 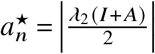, and combining Equation (12) with Equation (13), we get:

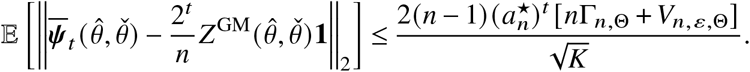

By Markov’s inequality and Equation (12), we get that for every *z* > 0

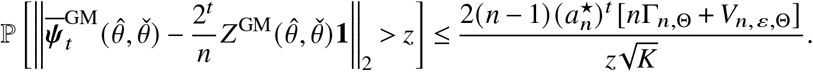

We let the RHS be equal to 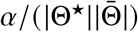, which corresponds to letting 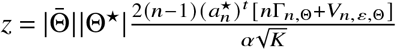. By applying a union bound over 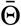 and 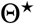 we have that for every 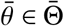 and 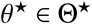, with probability at least 1 − *α*,

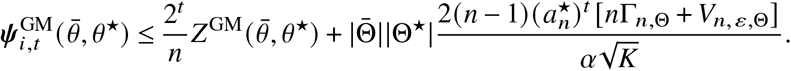

Conditioned on *F*^*c*^, the above becomes

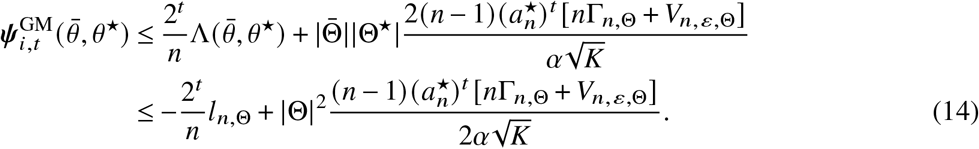

Note that we have used 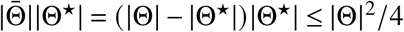. To make Equation (14) at most −*ϱ* for some *ϱ* > 0, we need to set

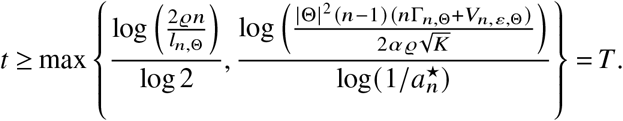

The log-belief ratio threshold implies that for all *θ*^★^ ∈ Θ^★^, 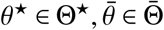 we have that 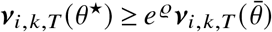. To determine the belief threshold *τ*, note that

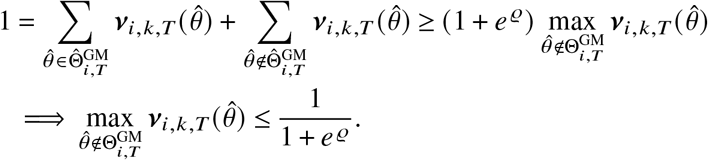

Moreover, we can prove that 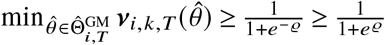 since *ϱ* > 0 which shows that any value in [1/(1 + *e* ^*ϱ*^), 1/(1 + *e*^− *ϱ*^)] is a valid threshold. This yields that 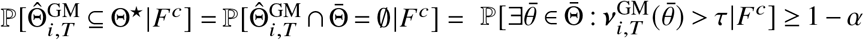. Subsequently, by letting 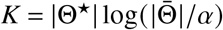 we get that

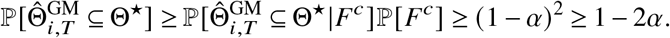

Finally, we show that 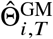 is non-empty with high probability. Conditioned on *F*^*c*^, we have that 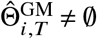 since at least one *θ*^★^ ∈ Θ^★^ satisfies 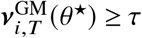. Using the law of total probability and the bound 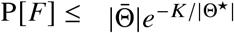, we get

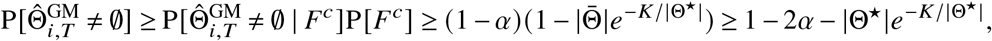

where the last step uses 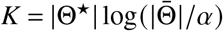, so 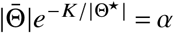.

#### AM estimator

We let 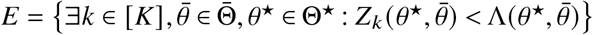. Using similar arguments to Theorem 2, we can deduce that 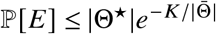. By setting 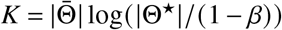, we make ℙ[*E*] ≤ 1 − *β*.

Conditioned on *E* ^*c*^ and by applying Markov’s inequality, similarly to Equation (14) for the GM estimator, we have that for all *θ*^★^ ∈ Θ^★^, 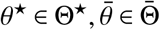 and run *k* ∈ [*K*]

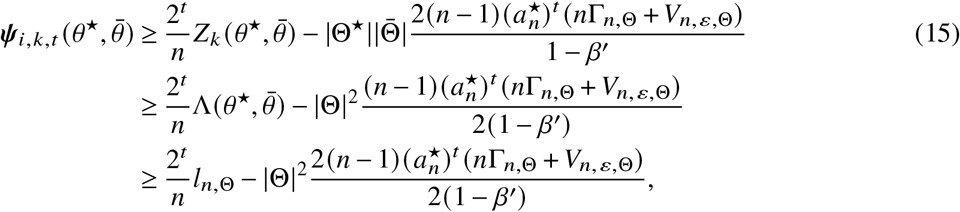

with probability *β*^′^ for some *β*^′^ ∈ (0, 1). To make the above at least *ϱ* for some *ϱ* > 0 it suffices to pick

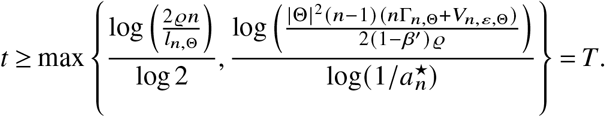

The log-belief ratio threshold implies that for all *θ*^★^ ∈ Θ^★^, 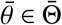 we have that 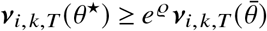. Similarly, any value in [1/(1 + *e* ^*ϱ*^), 1/(1 + *e*^− *ϱ*^)] is a valid threshold, we can show that

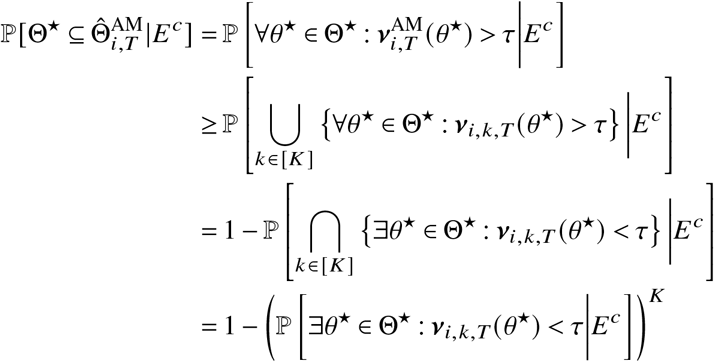

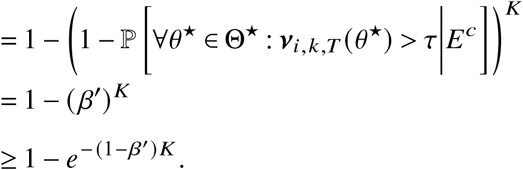

We set 1 − *β*^′^ = log(1/(1 − *β*))/*K* and have that

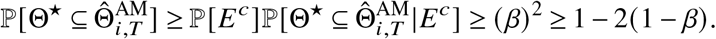

These finally yield

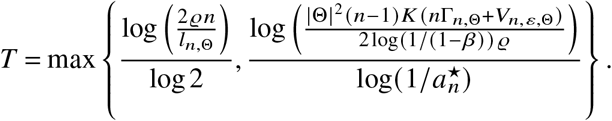

Similarly, we show that 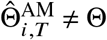 with high probability. Conditioned on *E* ^*c*^, at least one 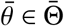 satisfies 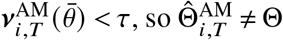. Using the law of total probability and the bound 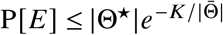, we get

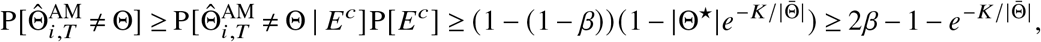

where the last step uses 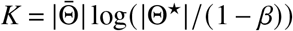, so 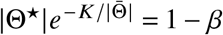.

#### Privacy (for both estimators)

Minimizing the convergence time *T* corresponds to minimizing *V*_*n,ε*,Θ_. Since *V*_*n,ε*,Θ_ is separable over the agents, it suffices to solve the problem of minimizing the variance of each noise variable independently.

If an adversary can eavesdrop only once and has access to any round *k* ∈ [*K*], and we have |Θ| states, by the composition theorem, the budget *ε* should be divided by *K* |Θ|. Thus, the problem of finding the optimal distribution D_*i*_ (*ε*) on ℝ, corresponds to the following optimization problem studied in Koufogiannis et al. (2015), Papachristou and Rahimian (2025), for all *k* ∈ [*K*]:

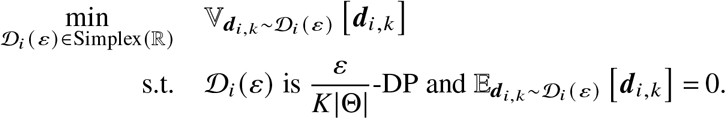

The optimal solution to this problem is given by Koufogiannis et al. (2015), Papachristou and Rahimian (2025). For privatizing beliefs in our problem, this corresponds to selecting 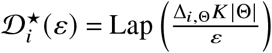 where Δ_*i*,Θ_ is the sensitivity of the log-likelihood of the *i*-th agent.

#### Differentially Private Outputs

Due to the postprocessing property of DP (see Proposition 2.1 of Dwork and Roth (2014)), the resulting estimates are *ε*-DP with respect to the private datasets.

Q.E.D.

### G.4. Proof of Theorem 3: Asymptotic Behavior of the Two-Threshold Algorithm

For simplicity, we define 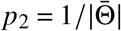 and *p*_1_ = 1 − 1/|Θ^★^|, and assume that *ϱ*^thres,1^ = *ϱ*^thres,2^ = *ϱ* which implies that 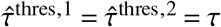 and 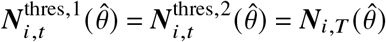.

#### Type I estimator 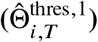

To determine the asymptotic Type I error probability *α* of 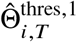, we assume that the threshold takes the form *τ*^thres,1^ = (1 + *π*_1_) *p*_1_. Then

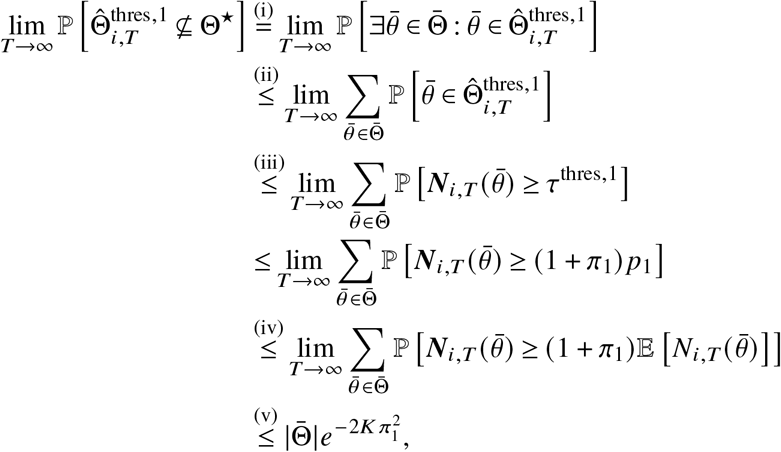

where the result is derived by applying (i) the definition of 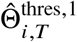, (ii) union bound, (iii) the definition of the threshold *τ*^thres,1^ = (1 + *π*_1_) *p*_1_, (iv) the fact that 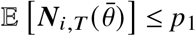 for all 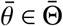 as *T* → ∞, and the Chernoff bound on 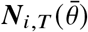. Therefore, to make the above *α*, it suffices to choose 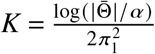.

#### **Type II estimator** 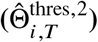

To determine the asymptotic Type II error probability 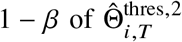, we assume that the threshold takes the form *τ*^thres,2^ = (1 − *π*_2_) *p*_2_. Then

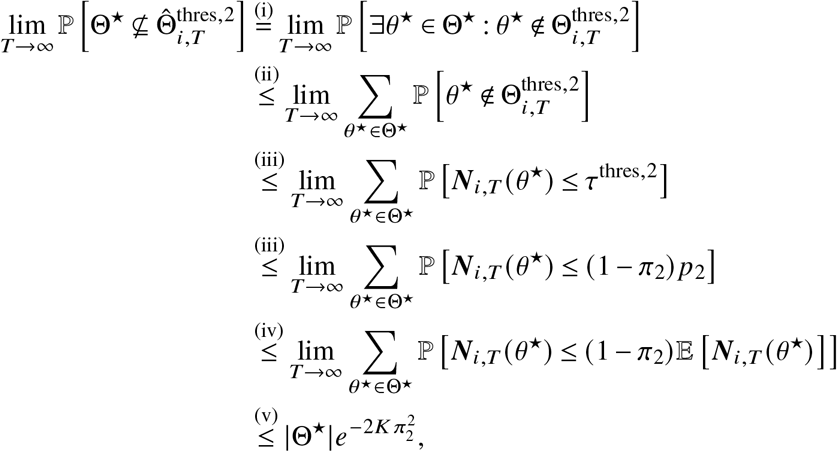

where the result is derived by applying (i) the definition of 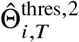, (ii) union bound, (iii) the definition of the threshold *τ*^thres,2^ = (1 − *π*_2_) *p*_2_, (iv) the fact that E [***N***_*i,T*_ (*θ*^★^)] ≥ *p*_2_ for all *θ*^★^ ∈ Θ^★^ as *T* → ∞, and (v) the Chernoff bound on 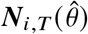. Therefore, to make the above less than 1 − *β*, it suffices to choose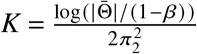.

#### Differentially Private Outputs

Due to the immunity to post-processing of DP (Dwork and Roth 2014, Proposition 2.1), the resulting estimates are *ε*-DP with respect to private datasets.

Q.E.D.

### G.5. Proof of Theorem 4: Finite-Time Guarantees for the Two-Threshold Algorithm

For simplicity, we assume that *ϱ*^thres,1^ = *ϱ*^thres,2^ = *ϱ* which implies that 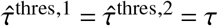 and 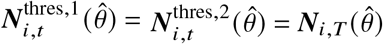.

#### Type I estimator

From the analysis of Theorem 2 (see Equation (14)), setting

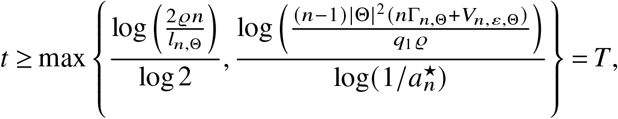

guarantees that 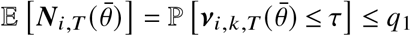 for all 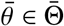. Then following the analysis similar to Theorem 4, using the union bound and the Chernoff bound, we can prove that 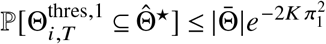. Subsequently, setting 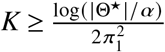 makes the error to be at most *α*.

#### Type II estimator

From the analysis of Theorem 2 (see Equation (15)), setting

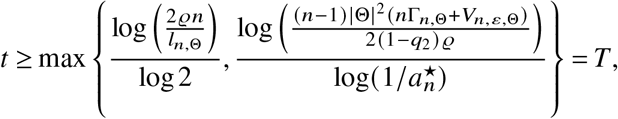

guarantees that E [***N***_*i,T*_ (*θ*^★^)] = ℙ [***ν***_*i,k,T*_ (*θ*^★^) > *τ*] ≥ *q*_2_ for all *θ*^★^ ∈ Θ^★^. Then following a similar analysis to Theorem 4 using the union bound and the Chernoff bound, we can prove that 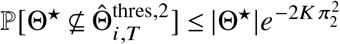. Subsequently, setting 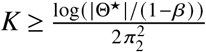 makes the error at most 1 − *β*.

#### Optimal Distributions

The proof is exactly the same as in Theorem 2.

#### Differentially Private Outputs

Due to the post-processing property of DP, see Dwork and Roth (2014, Proposition 2.1), the resulting estimates are *ε*-DP with respect to private datasets.

Q.E.D.

## Appendix H

**Proofs and Convergence Analysis for Private Distributed Hypothesis Testing**

### H.1. Proof of Proposition 1: Simple Hypothesis Testing at Significance Level *α*

Let 0 < *α*^′^, *α*^′′^ < 1, *α*^′^ + *α*^′′^ = *α* to be determined later. The centralized UMP test at level *α*^′′^ defines a threshold *ϱ*_*c*_ such that *θ* = 0 is rejected if 2Λ(1, 0) ≥ *ϱ*_*c*_. The threshold *ϱ*_*c*_ is selected such that ℙ[2Λ(1, 0) ≥ *ϱ*_*c*_ |*θ* = 0] = *α*^′′^. For the decentralized test, by the GM algorithm convergence analysis, we know that after *K* runs and *T* iterations given by the GM algorithm with Type I guarantee *α*^′^ and threshold *ϱ*^GM^ = 1 we have the following event

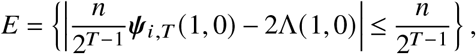

Thus, for these values of *T, K* we set *ϱ*_*d*_ = *ϱ*_*c*_ − 1. From the above relations, we get that under *θ* = 0 and *E*: {2Λ(1, 0) ≥ *ϱ*_*c*_} =⇒ *ψ*_*i,T*_ (1, 0) ≥ *ϱ*_*d*_ . This means that if the centralized test rejects, then the decentralized test also rejects *θ* = 0 with probability at least 1 − *α*^′^. The Type I error is:

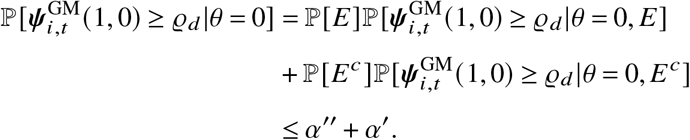

We choose *α*^′^ = *α*/2, *α*^′′^ = *α*/2 such that *α*^′^ + *α*^′′^ = *α*. The privacy guarantee is a direct consequence of DP’s post-processing property.

Q.E.D.

### H.2. Extension to Composite Hypotheses

The extension to composite hypotheses is straightforward since, if we consider the generalized likelihood ratio test and run the GM algorithm with the parameters of Proposition 1 we would obtain that for *T* and *K* set as in above, with probability at least 1 − *α*/2 the log-belief ratio statistic satisfies

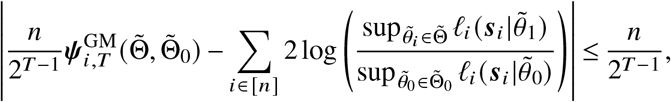

where the centralized generalized log-likelihood ratio statistic 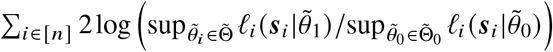 converges in distribution to 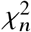 for sufficiently large *n*_*i*_. If 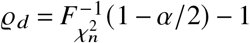, then the composite hypothesis test is *ε*-DP and has Type I error at most *α*.

## Appendix I

**Proofs and Convergence Analysis for Private Distributed Online Learning**

### I.1. Log-Belief Ratio Notations

Let 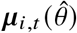 be the beliefs for the non-private system, and let 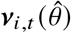 be the private estimates for agent *i* ∈ [*n*] and round *t* ≥ 0. For the non-private algorithm (Algorithm 4) all pairs 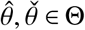 we let 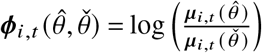 and 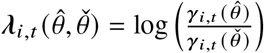 be the log belief ratios and the log-likelihood ratios respectively for agent *i* ∈ [*n*] round *t* ≥ 0. For the private algorithm (Algorithm 5) we let 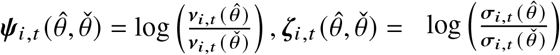, and 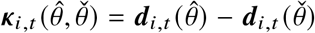 be the private log-belief ratio, log-likelihood ratio, and noise difference ratios respectively for agent *i* ∈ [*n*], and round *t* ≥ 0. We also let the vectorized versions 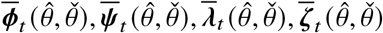 respectively, where the vectorization is over the agents *i* ∈ [*n*]. Finally, for each agent *i* ∈ [*n*] and pair of states 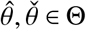 we define the KL divergence between the states 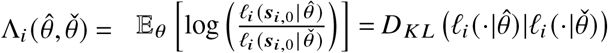.

### I.2. Auxiliary Lemmas

Before proving the main result for online learning, we prove the following lemma regarding the rate of convergence of the Césaro means.

#### Lemma 2.

*Let* 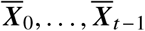 *be i*.*i*.*d. random variables (vectors), let A be an irreducible and aperiodic(primitive) doubly stochastic matrix with the second largest eigenvalue modulus* |*λ*_2_(*A*) | < 1, *and let* 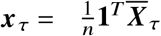 *for all* 0 ≤ *τ* ≤ *t* − 1, *with* 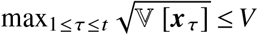. *Then*

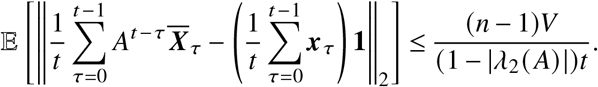

*Proof*. For 0 ≤ *τ* ≤ *t* − 1, we have

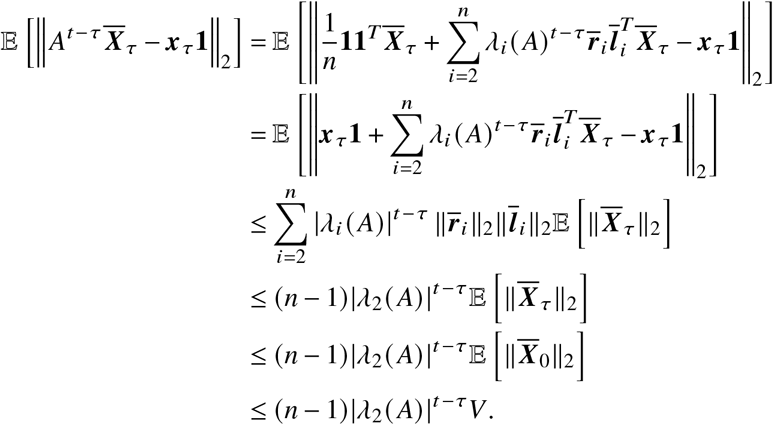

Therefore,

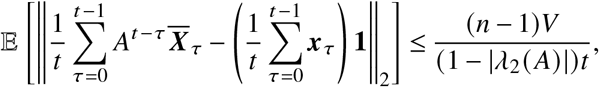

where we use 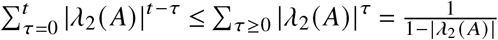 Q.E.D.

In the sequel, we show the main result for online learning.

### I.3. Proof of Theorem 5: Finite-Time Guarantees for Private, Distributed Online Learning

The dynamics of the log-belief ratio obey 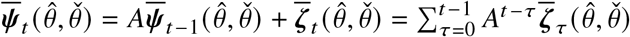, where 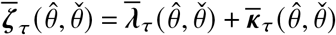. We apply Lemma 2, and get that

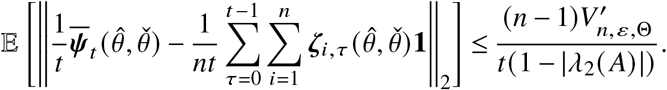

where,

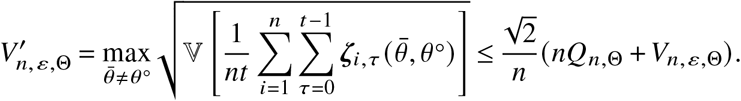

Moreover,

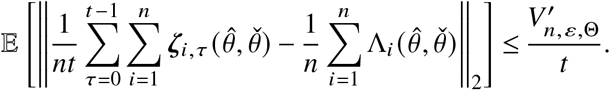

Therefore, by Markov’s inequality and the triangle inequality, we get that for every *z* > 0

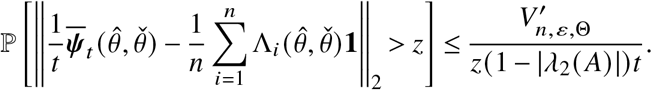

Therefore, we can show that by applying a union bound over Θ^★^, we have that with probability 1 − *η*, for all 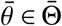

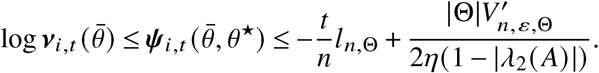

To make the RHS the log-belief threshold at most some value −*ϱ* for some *ϱ* > 0, we require

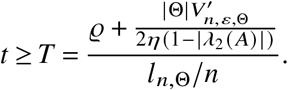

To determine *ϱ*, note that 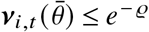for all 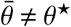, and, thus,

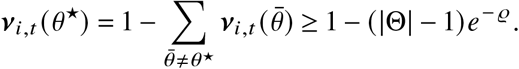

To determine a valid value of *ϱ*, we require 1 − (|Θ| − 1)*e*^− *ϱ*^ ≥ *e*^− *ϱ*^ which yields *ϱ* ≥ log(|Θ|). Therefore, setting *ϱ* = log(|Θ|) and subsequently

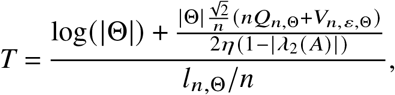

we get that 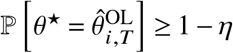.

#### Privacy

Similarly to Theorem 2, the optimal distributions are those that minimize variance subject to privacy constraints. Because there are |Θ| states, the budget *ε* should be divided by |Θ|, resulting in 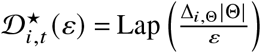. Q.E.D.

#### Differentially Private Outputs

Due to the post-processing property of DP (see Proposition 2.1 of Dwork and Roth (2014)), the resulting estimates are *ε*-DP with respect to the private datasets.

## Appendix J

**Differentially Private Distributed Survival Analysis for Clinical Trials**

### J.1. Sensitivity of the Proportional Hazards Model

The released statistic is,

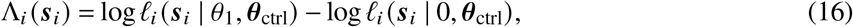

which tests *H*_0_ : *θ*_trt_ = 0 against *H*_1_ : *θ*_trt_ = *θ*_1_ with the controls ***θ***_ctrl_ held common to the two models.

First, we take ***x***_*i j*_ ∈ {0, 1}, |*θ*_1_| ≤ *B*_*θ*_, ∥ ***z***_*i j*_ ∥ ≤ *B*_*z*_, and 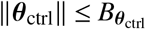. Using log 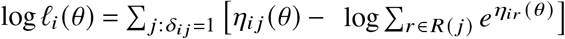with 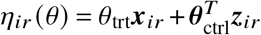and *R*(*j*) = { *j* ^′^ : ***t***_*i j*_′ ≥ ***t***_*i j*_ }. The numerator terms regarding the controls cancel between the two log-likelihoods and

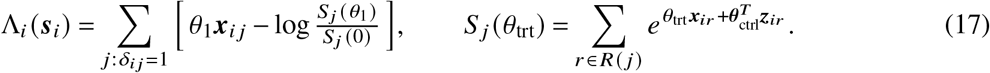

Second, because ***x***_*ir*_ ∈ {0, 1}, split the risk mass into untreated and treated parts, 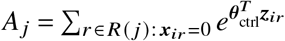and 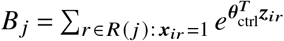, and let *π* _*j*_ = *B* _*j*_ /(*A* _*j*_ + *B* _*j*_) ∈ [0, 1] be the treated fraction of the risk set.

Then 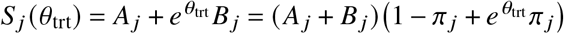, so log{*S* _*j*_ (*θ*_1_)/*S* _*j*_ (0)} = *φ*(*π* _*j*_) with

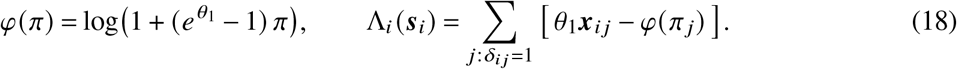

Each event contributes *λ* _*j*_ = *θ*_1_***x***_*i j*_ − *φ*(*π* _*j*_), which yields |*λ* _*j*_ | ≤ |*θ*_1_| ≤ *B*_*θ*_ .

Next, we let ***s***_*i*_ and 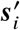 differ in patient *k*. We have two cases:

1. If *k* experiences an event, its own contribution *λ*_*k*_ is added or removed, contributing at most *B*_*θ*_ .
2. With weight 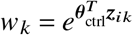, patient *k* belongs to the risk set of *every* event with ***t***_*i j*_ ≤ ***t***_*ik*_, adding *w*_*k*_ to *A* _*j*_ (if untreated) or *B* _*j*_ (if treated), thereby changing *φ*(*π* _*j*_). Since *φ* obeys 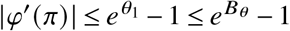, and removing *k* shifts the treated fraction by

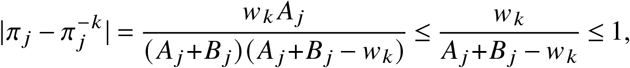

where 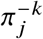is the treated fraction of the risk set without *k*. This yields

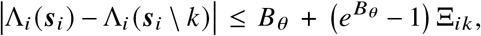

with

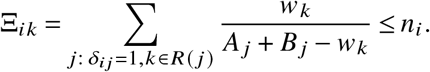

Therefore, the global sensitivity satisfies

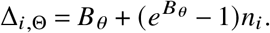

**Figure J.1.**
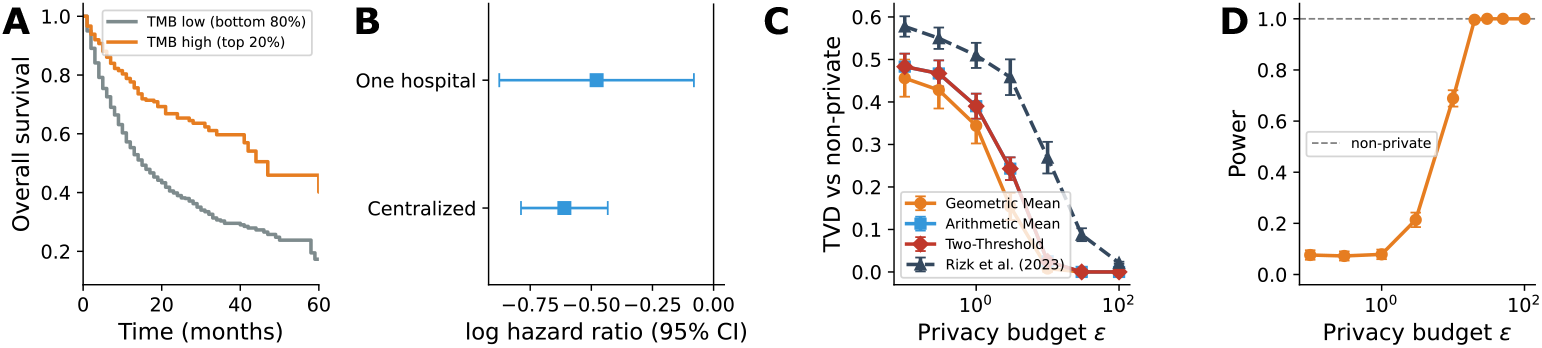
**Differentially private distributed survival analysis on the advanced-cancer cohort** of Samstein et al. (2019), testing the effect of high tumor mutational burden (top quintile) adjusted for age, sex, drug class, sequencing panel, and cancer type, and *n* = 5 fully connected centers. (A) Kaplan-Meier survival curves for the TMB-high and TMB-low groups. (B) TMB-high log hazard ratio with 95% confidence intervals, fitted centrally and at a single hospital. (C) Total variation distance between the private and non-private beliefs versus *ε*, together with the differentially private first-order method of Rizk et al. (2023) (D) Statistical power of the GM algorithm against *ε* at a significance level *α* = 0.05. The dashed line shows the non-private power. Because this cohort is larger and the TMB effect stronger than the ddI effect, the federated test reaches full power by *ε* ≈ 5, well below the budget required on ACTG 175 (Figure 2).

#### A Practical Mechanism

In practice, for a clipping threshold *C* > 0, we release 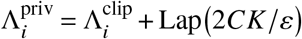, where 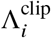is the magnitude-clipped statistic:

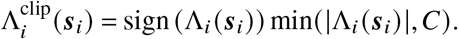

Since 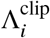takes values in [−*C, C*], its *ℓ*_1_-sensitivity is at most 2*C*, so the Laplace mechanism at scale 2*CK*/*ε* is *ε*-DP over the *K* rounds, and the subsequent belief exchange is also protected by post-processing immunity.

### J.2. Additional Experimental Results with the Cancer Dataset

Mirroring the adjusted analysis of Section 4, we replicate the pipeline on the advanced-cancer cohort of Samstein et al. (2019): *N* = 1,631 of the 1,662 patients treated with immune checkpoint inhibitors (excluding 31 with zero recorded survival time), with 810 deaths. The exposure is a binary indicator of high tumor mutational burden (TMB in the top quintile, the standard immunotherapy cutoff), and we adjust for age, sex, drug class (PD-1/PD-L1, CTLA-4, or combination), sequencing panel, and cancer type. Because TMB is measured on panel-specific assays and its distribution varies significantly across tumor types, the multicenter oncology study would have to account for them.

Figure J.1 summarizes the results. High TMB is strongly protective: the centralized adjusted analysis estimates a log hazard ratio of −0.61 (95% CI [−0.79, −0.43], *P* < 10^−10^), whereas a single hospital holding one fifth of the cohort recovers a consistent but far less certain estimate (−0.48, 95% CI [−0.88, −0.08]). The distributed differentially private belief exchange reproduces the centralized conclusion, with the privacy-utility gap closing as *ε* grows and the geometric-mean rule again converging fastest.

### J.3. Time and Space Complexity

We first start by analyzing the time complexity of our method. Specifically, in the distributed MLE setting, if agent *i* has *n*_*i*_ data points and model *ℓ*_*i*_ which can be computed in *T*_*i*_ (*n*_*i*_) for one value of *θ* ∈ Θ, then the distributed MLE is parallelizable between the agent and the total time complexity per agent is *O*(|Θ|*K T*_*i*_ (*n*_*i*_) +*T* deg_G_ (*i*))) which includes an initialization cost of |Θ|*K T*_*i*_ (*n*_*i*_) to calculate noisy likelihoods. For example, in the case of the distributed proportional hazards model, the likelihoods can be computed in *T*_*i*_ (*n*_*i*_) = *O* (*n*_*i*_ log *n*_*i*_). Then, there is a communication cost, which consists of the number of belief exchanges dictated by the communication complexity *KT*, and the cost of propagation for each iteration, which is O(|Θ |*K* deg_G_ (*i*)) for all states. Similarly, in the OL regime, the total time complexity per agent is 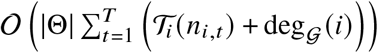. Finally, in terms of space complexity, each agent can simply maintain their belief in each state, giving a space complexity O(*K*|Θ|) during communication.

### J.4. Runtime Comparison with Homomorphic Encryption Methods

To further test the practical applicability of our method, we compare the runtime of our method with existing homomorphic encryption methods (HE) and, in particular, the FAMHE method introduced in Froelicher et al. (2021) as the number of providers (*n*) grows and the number of data grows. Both our method and FAMHE are capable of performing survival analysis in a distributed regime; however, they apply different privacy protections and cannot be compared prima facie. Generally, HE methods have the strongest possible privacy. However, these protections fall short in scalability considerations, for which DP with a small *ε* is a better alternative in terms of efficiency. Furthermore, the most recent HE methods designed for multicenter trials do not have open-source implementations, making direct comparison difficult (see, e.g. Froelicher et al. (2021), Geva et al. (2023)).

First, our aim is to compare the two methods in the same dataset and experiment, which corresponds to distributed survival analysis with cancer data introduced in Samstein et al. (2019). Specifically, Samstein et al. (2019) finds that tumor mutational burden (TMB) is a significant predictor of survival outcomes in patients with metastatic cancer undergoing immune checkpoint inhibitor (ICI) therapy. Analyzing data from 1,662 advanced cancer patients, the authors demonstrated that higher TMB levels are associated with better survival prospects.

Froelicher et al. (2021) use the data from Samstein et al. (2019) and perform survival analysis (see Figure 2 in their paper) by splitting the data equally among *n* providers. Then, the authors report the runtime as a function of *n* for three datasets: with 4096 time points (t.p.), 8192 t.p., and a dataset that consists of 10 times the original data, and report the total runtime in Figure 2(c). To construct the fairest possible comparison, we consider the same dataset and the distributed proportional hazards model with TMB as a covariate and two hypotheses, null (*θ* = 0) and a composite alternative (*θ* ≠ 0), similarly to our initial example for ACTG (see Section 1.1). The first two datasets (4096 and 8192 t.p.) are constructed by resampling the original data with replacement. We choose *ε* to be 0.1, which corresponds to a very strong privacy regime, significantly less than the privacy budgets used in the 2020 US Census (US Census 2020, Garfinkel 2022) and other healthcare contexts (Ficek et al. 2021, Dwork et al. 2019). Finally, we chose the error bounds to be no more than 0.1. In Figure J.2, we report the per-agent cost of inference for *n* ∈ {6, 12, 24, 48, 96} in a fully connected topology, analogous to Figure 2(c) of Froelicher et al. (2021). Our reported runtimes range from ∼ 10^−2^s to ∼ 1s, which is a 10 × −1000× improvement over the runtimes reported in Froelicher et al. (2021).

**Figure J.2.**
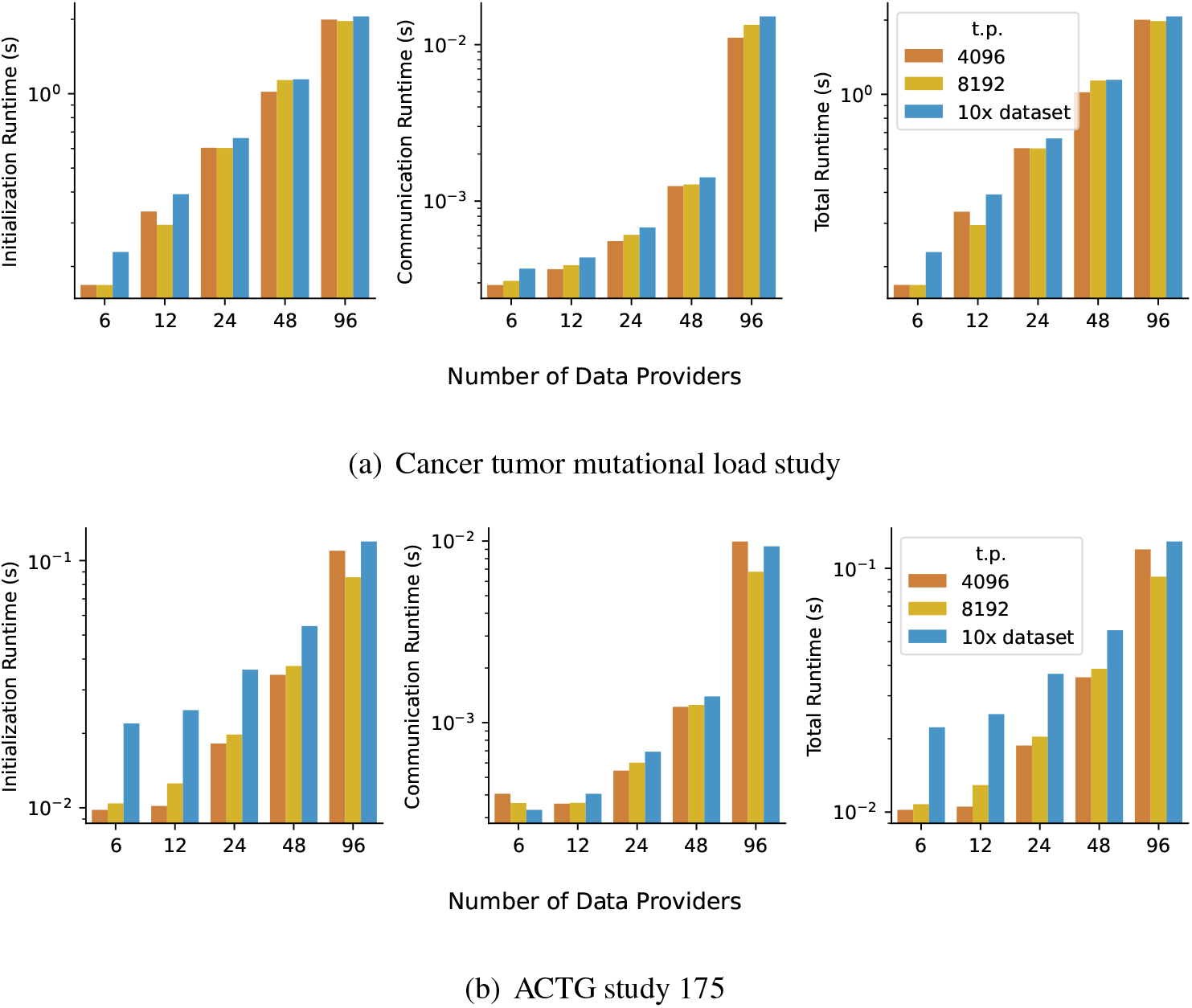
Runtime of distributed MLE algorithm for (a) the study by Samstein et al. (2019) on the top and (b) the ACTG study on the bottom, for varying values of *n* used in measuring the performance of the FAMHE method Froelicher et al. (2021). The privacy budget is set to *ε* = 1 (tight privacy), and the error rates are set to *α* = 1 − *β* = 0.05, and the thresholds are set to *ϱ*^AM^ = *ϱ*^GM^ = 0.1. In accordance with Froelicher et al. (2021), we have constructed 3 datasets: a dataset consisting of 4096 timepoints (t.p.) via resampling the original data with replacement, a dataset of 8192 t.p. via resampling the original data with replacement, and a dataset which consists of 10 times the original data.

### J.5. Comparison with First-order Methods

We compare with the first-order method of Rizk et al. (2023) with graph-homomorphic noise. The distributed optimization problem we are solving is

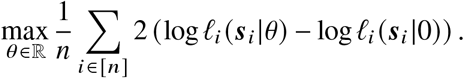

The distributed updates on the parameters with graph-homomorphic noise according to Rizk et al. (2023) are equivalent to

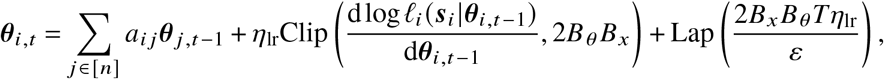

where *η*_lr_ is the learning rate, and the Clip operation clips the gradient to have *ℓ*_1_ norm at most 2*B*_*θ*_ *B*_*x*_. To derive the Laplace noise, note that the gradient decomposes over patients, 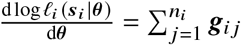with 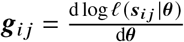, and each per-patient gradient satisfies ∥ ***g***_*i j*_ ∥_1_ ≤ *B*_*θ*_ *B*_*x*_ under ∥***x***_*i j*_ ∥ ≤ *B*_*x*_ and ∥***θ*** ∥ ≤ *B*_*θ*_ . Hence, two datasets differing by the addition or removal of one patient yield summed gradients that differ by at most *B*_*θ*_ *B*_*x*_ in *ℓ*_1_ norm, and because the *ℓ*_1_ clip is 2-Lipschitz, their clipped gradients differ by at most 2*B*_*θ*_ *B*_*x*_. The *ℓ*_1_-sensitivity of the update term *η*_lr_Clip(·) is therefore at most 2*η*_lr_*B*_*θ*_ *B*_*x*_. Additionally, since the per-agent budget is *ε*, we need to scale the budget by *T* . In the simulations, we use *B*_*θ*_ = 1, *η*_lr_ = 0.001, and in order to have a fair comparison to our algorithm, we set *T* to be equal to the number of iterations we run our algorithms. For large sample sizes the *P* values are given as 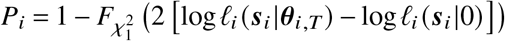.

## Appendix K

**Differentially Private Distributed Genetic Association Analysis**

This section provides the supporting details for the region study of Section 4.2, including the sensitivity analysis underlying the privacy calibration and additional experimental results.

### K.1. Setup and Notation

Let 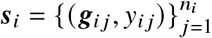denote the dataset at center *i*, where ***g***_*i j*_ ∈ {0, 1, 2}^*M*^ is the genotype vector and *y*_*i j*_ ∈ ℝ is the phenotype of individual *j*, generated under the linear model

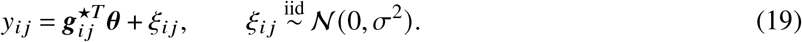

Let 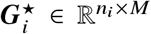denote the standardized genotype matrix with (*j, m*) entry 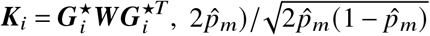, and let 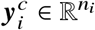denote the centered phenotype vector with entries 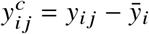. The weight matrix is ***W*** = diag(*w*_1_, …, *w*_*M*_) with weights *w*_*m*_ = Beta(MAF_*m*_; 1, 25)^2^, optionally clipped at a maximum value *w*_max_ ≤ 1. Define the *n*_*i*_ × *n*_*i*_ kernel matrix:

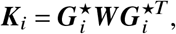

whose (*j, j* ^′^) entry 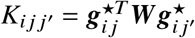is the weighted genetic similarity between individuals *j* and *j* ^′^. We employ the SKAT statistic (Wu et al. 2011):

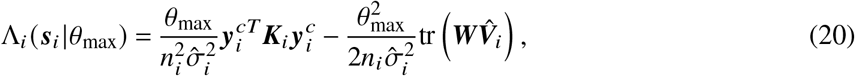

where 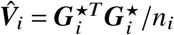is the empirical LD matrix and 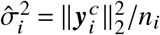.

The global *ℓ*_1_-sensitivity is:

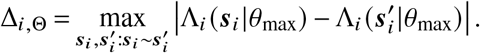

for datasets ***s***_*i*_, 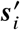such that they differ in at most one patient.

### K.2. Bounding the Sensitivity of the SKAT Statistic

#### Theorem 6.

*Under model* (19), *assume* |*θ*_max_| ≤ *B*_*θ*_ *and let δ* ∈ (0, 1) *be a failure probability. Then, with probability at least* 1 − *δ:*

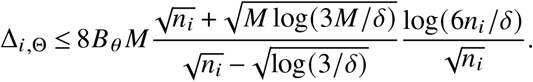

**Proof**. Let *δ* ∈ (0, 1). We apply a union bound to guarantee that three separate random events behave well simultaneously, allocating a failure probability of *δ*/3 to each.

First, because the phenotypes are drawn from a sub-Gaussian distribution, we can bound the maximum absolute value of the centered phenotype vector. With probability at least 1 − *δ*/3, the maximum centered phenotype is bounded by *C*_*σ*_, where:

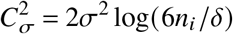

We denote this high-probability event as 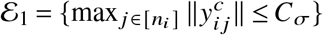.

Second, the SKAT statistic divides by the empirical variance 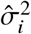. We must ensure this denominator does not become arbitrarily small. Because 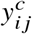are sub-Gaussian, 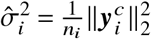, the standard sub-exponential tail bounds yields that, with probability at least 1 − *δ*/3:

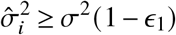

where 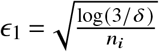. We denote this event as ℰ_2_.

Third, we apply the Matrix Bernstein inequality to bound the spectral gap between the empirical covariance matrix 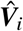and the population correlation matrix ***R***. With probability at least 1 − *δ*/3:

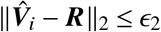

where 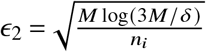. By Weyl’s inequality, we can bound the maximum eigenvalue of the weighted matrix:

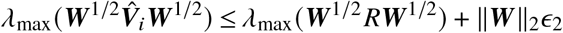

Because ***R*** is a correlation matrix (diagonals equal to 1), *λ*_max_(***W***^1/2^ ***RW***^1/2^) ≤ tr(***W R***) = tr(***W***) ≤ *M*. Furthermore, because ***W*** is diagonal with positive weights, ∥***W***∥_2_ ≤ tr(***W***) ≤ *M*. Therefore:

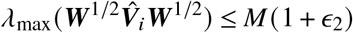

We denote this event as ℰ_3_. By the union bound, the intersection event ℰ = ℰ_1_ ∩ ℰ_2_ ∩ ℰ_3_ holds with probability at least 1 − *δ*.

Next, we evaluate the sensitivity between two adjacent datasets, ***s***_*i*_ and 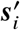, differing only by individual *k*. We decompose the centered phenotype vector as 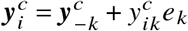.

Expanding the quadratic form of the SKAT statistic for ***s***_*i*_ and 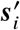and taking the difference 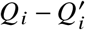, the terms not involving *k* cancel out, leaving a cross term (*A*_1_) and a squared term (*A*_2_):

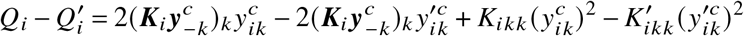

Conditioned on ℰ, we apply the Cauchy-Schwarz inequality and spectral identities to bound (*A*_1_):

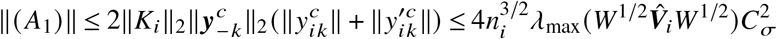

Using the triangle inequality and bounding the diagonal entries by the trace *K*_*ikk*_ ≤ tr(***W***), we bound (*A*_2_):

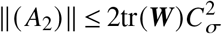

The trace term changes by at most 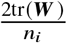, contributing 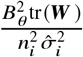to the sensitivity. For *n*_*i*_ large enough such that 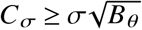this is bounded by 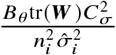.

Finally, dividing the quadratic bounds by the denominator 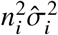and combining with the trace term gives the intermediate bound:

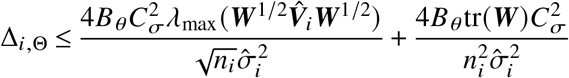

We absorb the second term, as it decays at a faster rate 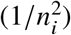and is negligible relative to the 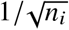term. We now substitute our finite-sample bounds from events ℰ_1_, ℰ_2_, and ℰ_3_:

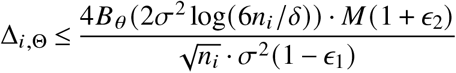

where

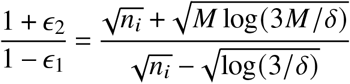

Combining everything together, we get

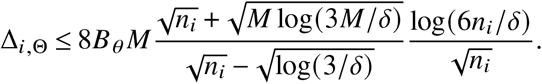

Q.E.D.

### K.3. A Practical Mechanism

The sensitivity above had polynomial dependence on the number *M* of SNPs. By applying clipping we can remove this dependence and achieve similar performance. Specifically, we privatize the SKAT statistic with the Laplace mechanism. The construction clips each individual’s contribution and calibrates the noise accordingly.

Write 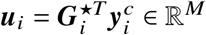for the weighted score vector of center *i*, so that the quadratic form *Q*_*i*_ is the weighted squared norm 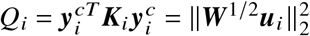and the statistic Λ_*i*_ of (20) is an affine function of *Q*_*i*_. The vector ***u***_*i*_ decomposes over individuals as 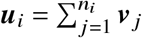with 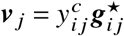, each term depending only on individual *j* . We clip every contribution in the *L*_1_ norm, 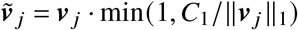 for a threshold *C*_1_ > 0, form 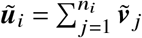, and release 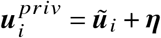with independent Laplace coordinates *η*_*m*_ ∼ Lap(2*C*_1_*K*/*ε*).

#### Theorem 7.

*Let C*_1_ > 0. *Releasing* 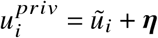*with η*_*m*_ ∼ Lap(2*C*_1_*K*/*ε*) *i*.*i*.*d. satisfies ε-DP over the K rounds. The SKAT statistic* 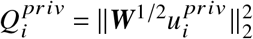*and* 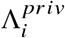*are ε-DP by post-processing*.

**Proof**. Because *v* _*j*_ depends only on individual *j*, replacing one individual leaves the other *n*_*i*_ − 1 clipped terms unchanged, so 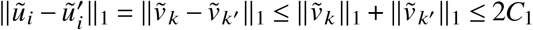by the triangle inequality and the clip. The *ℓ*_1_-sensitivity of *ũ*_*i*_ is therefore Δ_1_ = 2*C*_1_, and the Laplace mechanism with the stated scale is *ε*-DP. Post-processing gives the claim.

Q.E.D.

### K.4. Algorithm Performance for Different Configurations of Parameters

We perform sensitivity analysis for the genetic association analysis study described in Section 4.2 using the same parameters as in the main text. We keep some of these parameters fixed and vary others to show how the performance of our method changes.

**Figure K.1.**
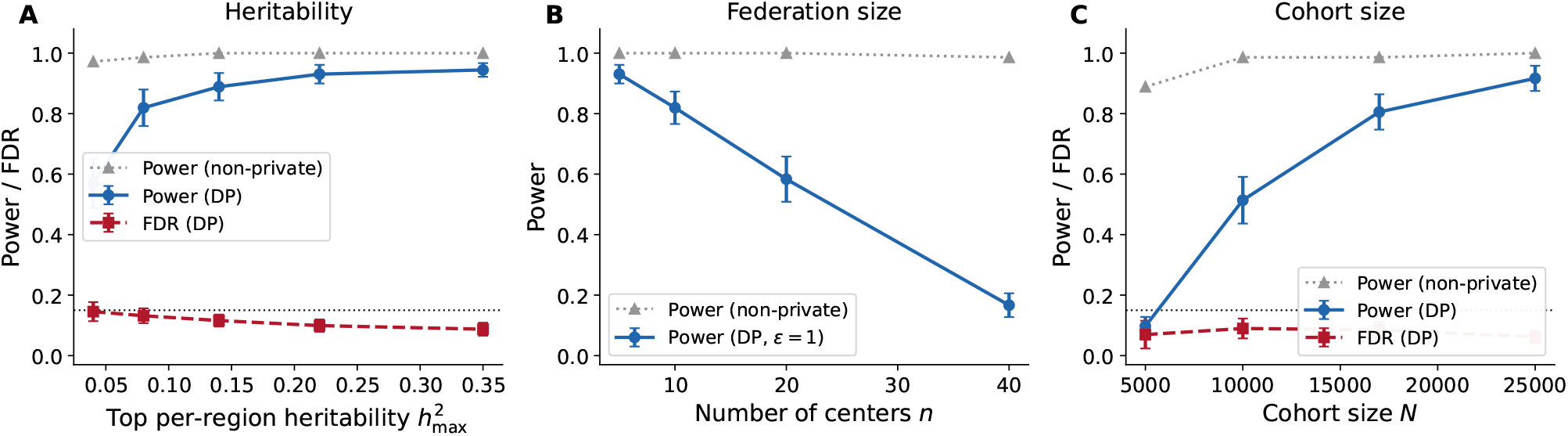
Sensitivity analysis of the genetic association analysis **(A)** Power and FDR versus per-region heritability. **(B)** Power versus the number of federating centers *n* at fixed total cohort. **(C)** Power and FDR versus cohort size *N*. When not varied, the parameters set as in Section 4.2.

Figure K.1A shows how power changes under varying heritability for each region, demonstrating that our algorithm achieves higher power with higher heritability, as expected.

Figure K.1B fixes the total cohort and varies *n* ∈ {5, 10, 20, 40} centers. As *n* grows, each center holds fewer individuals, and the mechanism injects Laplace noise at more sites, so power declines.

Figure K.1C shows power and FDR as a function of the cohort size *N* under a complete network. Power rises toward the non-private baseline as *N* grows, and the FDR is controlled at the nominal level for adequately sized cohorts.

### K.5. Additional Privacy-utility Curves

Figure K.2 verifies that the privacy-utility comparison of Figure 3 is robust to the choice of network topology: the organizational-level advantage persists when the organizations are arranged in a star network, and when the hospital-level network is taken to be fully connected.

**Figure K.2.**
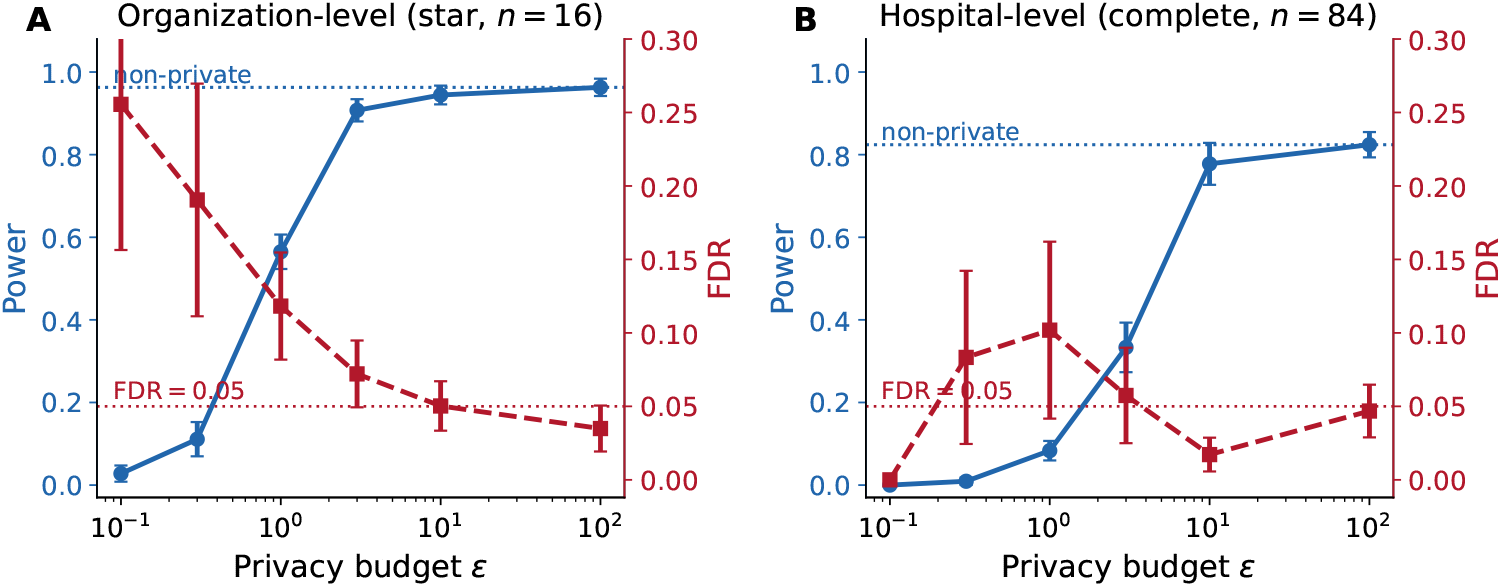
Robustness of the privacy utility curves of Figure 3 to the network topology. Panel (A) reproduces the same privacyutility curve with Figure 3A when the organizations are arranged in a star network with the central node being the biggest organization. Panel (B) reproduces the same privacy-utility curve with Figure 3B when the hospital-level network is fully connected. We have used *T* = 10, *K* = 2 and 10 independent runs.

## References

Abbas AE (2009) A Kullback-Leibler view of linear and log-linear pools. Decision Analysis 6(1):25–37.

ARPA-H (2024) ARPA-H Launches Groundbreaking Funding Opportunity to Improve Clinical Trials. https://arpa-h.gov/news-and-events/arpa-h-launches-groundbreaking-funding-opportunity-improve-clinical-trials, accessed: 2024-12-13.

Baumdicker F, Bisschop G, Goldstein D, Gower G, Ragsdale AP, Tsambos G, Zhu S, Eldon B, Ellerman EC, Galloway JG, et al. (2022) Efficient ancestry and mutation simulation with msprime 1.0. Genetics 220(3):iyab229.

Bullo F, Cortés J, Martinez S (2009) Distributed control of robotic networks: a mathematical approach to motion coordination algorithms, volume 27 (Princeton University Press).

Campbell TB, Smeaton LM, Kumarasamy N, Flanigan T, Klingman KL, Firnhaber C, Grinsztejn B, Hosseinipour MC, Kumwenda J, Lalloo U, et al. (2012) Efficacy and safety of three antiretroviral regimens for initial treatment of HIV-1: a randomized clinical trial in diverse multinational settings. PLoS Medicine 9(8):e1001290, URL http://dx.doi.org/10.1371/journal.pmed.1001290.

Casella G, Berger RL (2002) Statistical inference, volume 2 (Duxbury Pacific Grove, CA).

Clarke N, Vale G, Reeves EP, Kirwan M, Smith D, Farrell M, Hurl G, McElvaney NG (2019) GDPR: An impediment to research? Irish Journal of Medical Science 188:1129–1135.

Clemen RT, Winkler RL (1999) Combining probability distributions from experts in risk analysis. Risk analysis 19(2):187–203.

Cox DR (1972) Regression models and life-tables. Journal of the Royal Statistical Society: Series B (Methodological) 34(2):187–202.

Cyffers E, Bellet A, Upadhyay J (2024) Differentially private decentralized learning with random walks. Proceedings of the 41st International Conference on Machine Learning (ICML), volume 235 of Proceedings of Machine Learning Research (PMLR).

Davidson-Pilon C (2019) lifelines: survival analysis in Python. Journal of Open Source Software 4(40):1317.

DeGroot MH (1974) Reaching a consensus. Journal of American Statistical Association 69:118–121.

Dwork C, Kohli N, Mulligan D (2019) Differential privacy in practice: Expose your epsilons! Journal of Privacy and Confidentiality 9(2).

Dwork C, Roth A (2014) The algorithmic foundations of differential privacy. Foundations and Trends® in Theoretical Computer Science 9(3–4):211–407.

Erlich Y, Narayanan A (2014) Routes for breaching and protecting genetic privacy. Nature Reviews Genetics 15(6):409–421.

Ficek J, Wang W, Chen H, Dagne G, Daley E (2021) Differential privacy in health research: A scoping review. Journal of the American Medical Informatics Association 28(10):2269–2276.

Freund Y, Schapire RE (1997) A decision-theoretic generalization of on-line learning and an application to boosting. Journal of computer and system sciences 55(1):119–139.

Froelicher D, Troncoso-Pastoriza JR, Raisaro JL, Cuendet MA, Sousa JS, Cho H, Berger B, Fellay J, Hubaux JP (2021) Truly privacy-preserving federated analytics for precision medicine with multiparty homomorphic encryption. Nature communications 12(1):5910.

Garfinkel S (2022) Differential privacy and the 2020 US census. MIT Case Studies in Social and Ethical Responsibilities of Computing URL http://dx.doi.org/10.21428/2c646de5.7ec6ab93, winter 2022 issue.

Geva R, Gusev A, Polyakov Y, Liram L, Rosolio O, Alexandru A, Genise N, Blatt M, Duchin Z, Waissengrin B, et al. (2023) Collaborative privacy-preserving analysis of oncological data using multiparty homomorphic encryption. Proceedings of the National Academy of Sciences 120(33):e2304415120.

Gohari P, Wu B, Hawkins C, Hale M, Topcu U (2021) Differential privacy on the unit simplex via the dirichlet mechanism. IEEE Transactions on Information Forensics and Security 16:2326–2340.

Golub B, Sadler E (2016) Learning in social networks. Bramoullé Y, Galeotti A, Rogers B, eds., The Oxford Handbook on the Economics of Networks (Oxford University Press).

Hammer SM, Katzenstein DA, Hughes MD, Gundacker H, Schooley RT, Haubrich RH, Henry WK, Lederman MM, Phair JP, Niu M, et al. (1996) A trial comparing nucleoside monotherapy with combination therapy in HIV-infected adults with CD4 cell counts from 200 to 500 per cubic millimeter. New England Journal of Medicine 335(15):1081–1090.

He Z, Le Guen Y, Liu L, Lee J, Ma S, Yang AC, et al. (2021) Genome-wide analysis of common and rare variants via multiple knockoffs at biobank scale, with an application to Alzheimer disease genetics. The American Journal of Human Genetics 108(12):2336–2353, URL http://dx.doi.org/10.1016/j.ajhg.2021.10.009.

Homeland Infrastructure Foundation-Level Data (HIFLD) (2020) US hospital locations. Kaggle. Derived from the Homeland Infrastructure Foundation-Level Data (HIFLD), U.S. Department of Homeland Security. https://www.kaggle.com/datasets/andrewmvd/us-hospital-locations.

Homer N, Szelinger S, Redman M, Duggan D, Tembe W, Muehling J, Pearson JV, Stephan DA, Nelson SF, Craig DW (2008) Resolving individuals contributing trace amounts of DNA to highly complex mixtures using high-density SNP genotyping microarrays. PLoS genetics 4(8):e1000167.

Institute of Medicine (2009) Beyond the HIPAA Privacy Rule: Enhancing Privacy, Improving Health Through Research (Washington, DC: National Academies Press), s. J. Nass, L. A. Levit, and L. O. Gostin, eds.

Jackson MO (2008) Social and Economic Networks (Princeton, NJ: Princeton University Press).

Kao JL, Kang S (2026) Data privacy regulation and innovation. Available at SSRN 6104788 .

Kaplan EL, Meier P (1958) Nonparametric estimation from incomplete observations. Journal of the American statistical association 53(282):457–481.

Kayaalp M, Inan Y, Telatar E, Sayed AH (2024) On the arithmetic and geometric fusion of beliefs for distributed inference. IEEE Transactions on Automatic Control 69(4):2265–2280.

Koufogiannis F, Han S, Pappas GJ (2015) Optimality of the Laplace mechanism in differential privacy. arXiv preprint arXiv:1504.00065 .

Lalitha A, Javidi T, Sarwate AD (2018) Social learning and distributed hypothesis testing. IEEE Transactions on Information Theory 64(9):6161–6179.

Marden JR, Shamma JS (2012) Revisiting log-linear learning: Asynchrony, completeness and payoff-based implementation. Games and Economic Behavior 75(2):788–808.

National Institute of Allergy and Infectious Diseases (1995) A randomized, double-blind phase II/III trial of monotherapy vs. combination therapy with nucleoside analogs in HIV-infected persons with CD4 cells of 200-500/mm3. https://clinicaltrials.gov/study/NCT00000625, accessed: 2024-12-13.

Ness RB (2007) Influence of the HIPAA privacy rule on health research. JAMA 298(18):2164–2170.

Nissim K, Raskhodnikova S, Smith A (2007) Smooth sensitivity and sampling in private data analysis. Proceedings of the thirty-ninth annual ACM symposium on Theory of computing, 75–84.

Nyholt DR, Yu CE, Visscher PM (2009) On Jim Watson’s APOE status: genetic information is hard to hide. European Journal of Human Genetics 17(2):147–149.

Olshevsky A (2017) Linear time average consensus and distributed optimization on fixed graphs. SIAM Journal on Control and Optimization 55(6):3990–4014.

Papachristou M, Rahimian MA (2025) Differentially private distributed estimation and learning. IISE Transactions 57(7):756–772, URL http://dx.doi.org/10.1080/24725854.2024.2337068.

Rahimian MA, Jadbabaie A (2016) Distributed estimation and learning over heterogeneous networks. Communication, Control, and Computing (Allerton), 2016 54th Annual Allerton Conference on, 1314–1321 (IEEE).

Rizk E, Vlaski S, Sayed AH (2023) Enforcing privacy in distributed learning with performance guarantees. IEEE Transactions on Signal Processing 71:3385–3398.

Samstein RM, Lee CH, Shoushtari AN, Hellmann MD, Shen R, Janjigian YY, Barron DA, Zehir A, Jordan EJ, Omuro A, et al. (2019) Tumor mutational load predicts survival after immunotherapy across multiple cancer types. Nature genetics 51(2):202–206.

Sudlow C, Gallacher J, Allen N, Beral V, Burton P, Danesh J, Downey P, Elliott P, Green J, Landray M, et al. (2015) UK Biobank: an open access resource for identifying the causes of a wide range of complex diseases of middle and old age. PLoS medicine 12(3):e1001779.

US Census (2020) 2020 decennial census: Processing the count: Disclosure avoidance modernization. https://www.census.gov/programs-surveys/decennial-census/decade/2020/planning-management/process/disclosure-avoidance.html, accessed: 2023-05-18.

Wu MC, Lee S, Cai T, Li Y, Boehnke M, Lin X (2011) Rare-variant association testing for sequencing data with the sequence kernel association test. The American Journal of Human Genetics 89(1):82–93.

Zerhouni EA, Nabel EG (2008) Protecting aggregate genomic data. Science 322(5898):44, URL http://dx.doi.org/10.1126/science.1165490.

